# Comparative studies of genetic and phenotypic associations for 2,168 plasma proteins measured by two affinity-based platforms in 4,000 Chinese adults

**DOI:** 10.1101/2023.12.01.23299236

**Authors:** Baihan Wang, Alfred Pozarickij, Mohsen Mazidi, Neil Wright, Pang Yao, Saredo Said, Andri Iona, Christiana Kartsonaki, Hannah Fry, Kuang Lin, Yiping Chen, Huaidong Du, Daniel Avery, Dan Valle Schmidt, Canqing Yu, Dianjianyi Sun, Jun Lv, Michael Hill, Liming Li, Derrick A Bennett, Rory Collins, Robin G Walters, Robert Clarke, Iona Y Millwood, Zhengming Chen, China Kadoorie Biobank Collaborative Group

## Abstract

Proteomics offers unique insights into human biology and drug development, but few studies have directly compared the utility of different proteomics platforms. We measured 2,168 plasma proteins in 3,976 Chinese adults using both OLINK and SomaScan platforms and compared their genetic determinants and associations with traits and disease risk. For 1,694 proteins with one-to-one matched reagents, there was a modest between platform correlation (median rho=0.20). OLINK-proteins had fewer *trans-*pQTLs (766 vs 812 proteins) but more *cis-*pQTLs (725 vs 565) than SomaScan-proteins, including 342 with colocalising *cis*-pQTLs. Moreover, 1,095 OLINK- and 963 SomaScan-proteins showed significant associations with BMI, while 279 and 165 proteins were significantly associated with IHD, respectively. Addition of these IHD-associated proteins to conventional risk factors yielded NRIs for IHD of 15.3% and 17.1% for OLINK and SomaScan respectively. The results demonstrate the complementarity of different proteomic platforms and should inform assay selection in future population and clinical studies.

## Introduction

Proteins play a key role in human health and most drugs target proteins, including enzymes, antibodies, transport, or structural proteins. Measurements of plasma levels of proteins, particularly when combined with genetic and phenotypic information, can help to understand biological mechanisms and disease aetiology, improve disease risk prediction, and evaluate novel protein targets for drug treatment of specific diseases. Advances in high-throughput proteomic assays now enable measurement of thousands of plasma proteins, and their application in population and clinical studies is likely to transform the development of precision medicine.

The relative or absolute plasma levels of proteins can be measured using different technologies, including mass spectrometry and affinity-based methods. Mass spectrometry methods identify proteins based on peptides following enzymatic digestion and have been widely used for both targeted (focusing on a few hundred pre-defined proteins) and non-targeted (measuring up to 4,500 plasma proteins) approaches in clinical and population studies.^1–3^ However, due to the extensive pre-fractionation required,^4^ application of mass spectrometry in large-scale population studies remains challenging. In contrast, advances in high-throughput affinity-based protein profiling technology using the antibody-based OLINK^5^ or the aptamer-based SomaScan^6–8^ platform have made it possible to measure plasma levels of several thousand different protein markers simultaneously in large numbers of samples. The OLINK platform measures protein levels using paired antibodies binding a single target protein,^5^ while SomaScan employs slow off-rate modified aptamers (SOMAmers) as protein-binding reagents.^6–8^

Both OLINK and SomaScan platforms have recently been used in large-scale population studies, helping to identify genetic variants associated with plasma protein levels (i.e. pQTLs) and biomarkers of traits (e.g. BMI), diseases, and their progression.^9–12^ The latest OLINK and SomaScan platforms now include >5,000 and >11,000 protein reagents, respectively, targeting a large number of overlapping proteins. A few studies have recently compared, directly or indirectly, the analytical performance of different platforms and generally showed only modest correlations between protein levels measured by OLINK and SomaScan.^13–17^ Moreover, the protein-phenotype associations and pQTLs identified in these studies have also varied between platforms.^15–17^ In a recent large study, the findings were based mainly on indirect comparisons of different individuals enrolled in the UK Biobank or the deCODE study.^17^ Most studies that compared the platforms directly within the same blood samples were constrained by small sample sizes, limited numbers of overlapping proteins often only of high abundance, involvement of primarily European ancestry populations, and lack of in-depth genetic or concomitant analyses of proteins with phenotypes.^13–17^

We present direct comparative analyses of 2,168 proteins measured by both the OLINK Explore and SomaScan4.1 assays in ∼4,000 participants from an ischaemic heart disease (IHD) case-subcohort study in the China Kadoorie Biobank (CKB). The main aims of this study were to: (i) examine the correlations between plasma protein levels measured by the two platforms; (ii) compare the pQTLs identified in genome-wide association studies (GWAS); (iii) compare the associations of proteins with different traits (e.g. adiposity) and disease risks (e.g. IHD); and (iv) compare the performance of proteins for prediction of IHD risk, separately and in combination with conventional risk factors.

## Methods

### Study population and design

The CKB is a prospective cohort study with >512,000 adults recruited during 2004- 08 from 10 geographically diverse areas.^18^ At baseline and subsequent periodic resurveys of a random subset of participants, detailed data were collected from participants using laptop-based questionnaires (e.g. socio-demographic characteristics, medical history, and lifestyle habits) and physical measurements (e.g. anthropometry, blood pressure, heart rate, lung function). Non-fasting (with time since the last meal recorded) blood samples were collected, processed, aliquoted, and then stored in liquid nitrogen. After the initial baseline survey, the long-term health of the participants was monitored by linkage with local death or disease registries and with the national health insurance systems that record any episodes of hospitalisation.^18^ The study was approved by international, national, and local ethics committees, and all participants provided written informed consent.

The present analysis involved a case-subcohort study of 3,977 unrelated participants (1,951 incident IHD cases and 2,026 subcohort participants) who were genotyped, and had no prior history of cardiovascular diseases.^19^

### Genotyping

A total of 100,706 CKB participants were genotyped using a custom Affymetrix array, with 531,565 variants passing quality control (QC). They were converted to genome build 38 using CrossMap v0.6.1^20^ and checked for consistency by reversing the process (liftUnder). Variants were excluded if they were not mapped, mapped to different chromosomes, or not mapped back to the same locations after liftUnder, leaving 531,542 remaining variants for further analyses. They were pre-phased using SHAPEIT v4.2 (SHAPEIT v2.904 for chromosome X)^21^ and uploaded to the TOPMed^22^ or Westlake Biobank for Chinese^23^ server for imputation. Two sets of the imputed data were merged by selecting the imputed genotype with a higher imputation INFO score for each variant. Variants with an INFO score<0.3 or a minor allele frequency (MAF)=0 were excluded. Details of the genotyping and QC procedures have been previously described.^24^

### Proteomic assays

For the OLINK Explore 3072 assay, stored baseline plasma samples for 3,977 participants were retrieved from liquid nitrogen and thawed, and 40µl plasma was aliquoted into 96-well plates (including 8 wells per plate for external QC samples). Plates were shipped in two batches for assay at OLINK Laboratories (first batch, 1472 proteins in Uppsala, Sweden; and second batch, 1469 proteins in Boston, USA). Protein levels were normalised based on inter-plate controls and transformed using a pre-determined correction factor. The limit of detection (LOD) for reagents was defined using external QC samples. QC warnings for participant samples and assay warnings for plates were flagged based on deviations in the QC samples.^25^ Protein levels were provided in the arbitrary Normalized Protein eXpression (NPX) unit on a log2 scale. Among a total of 2,941 protein reagents, six were replicated across all four panels, resulting in 2,923 unique reagents. Only one measure for each duplicated OLINK reagent was used for the comparative analysis, as replicated protein levels had high correlations between panels (r>0.8).

For the SomaScan v4.1 assay, 60µl plasma aliquots in 2D-barcoded microtubes for the same 3,977 participants were sent to the Somalogic Laboratory in Colorado, USA for profiling by SomaScan Assay v4.1, which covers a total of 7,596 SOMAmers, including 7,335 targeting human proteins. Samples were randomised at the Somalogic laboratory and aliquoted into 96-well plates (11 wells allocated for external control samples, including 5 calibrator, 3 QC, and 3 buffer samples). One subcohort participant was excluded due to insufficient sample volume. For 91 participants with higher sample volumes, samples were split and run in duplicate. Only one measure from each duplicated sample was used for the comparative analyses as protein levels between duplicate samples were highly correlated (median rho>0.8). The raw SomaScan assay results were standardised based on external control samples to control for variability in microarrays and variation within and across plates. This also included an optional step of adaptive normalization by maximum likelihood (ANML) to an external reference to control for inter-sample variability.^26^ The final SomaScan data were supplied in both ANML and non-ANML versions in relative fluorescence units (RFU), which were further log-transformed (natural log) in the main analyses. The LOD for SOMAmers was defined using external buffer samples. QC checks were performed by comparing the median of QC samples on each plate to the reference, and a cross-plate QC check measure (pass/flag) was assigned to each SOMAmer.

### Protein target mapping and reagent matching

We mapped OLINK reagents and SOMAmers to proteins based on their UniProt^27^ IDs provided by OLINK and SomaScan.^27^ If two reagents from different platforms mapped to the same UniProt ID, we considered them to be an OLINK-SomaScan reagent pair. The main analysis focused on reagent pairs where one unique OLINK reagent was matched to one unique SOMAmer. Separate analyses were also undertaken to investigate proteins for which single SOMAmers matched to multiple OLINK reagents and single OLINK reagents matched to multiple SOMAmers.

### Statistical analyses

We calculated Spearman’s and Pearson’s correlation coefficients for each OLINK-SomaScan reagent pair, using both ANML and non-ANML SomaScan data. We annotated all OLINK-SomaScan reagent pairs in four areas, including: (i) OLINK or SomaScan assay-related factors (e.g. batch, dilution factor); (ii) OLINK or SomaScan data-related measures (e.g. % outliers [> 4 SD from the mean], % below LOD); (iii) protein characteristics retrieved from the UniProt Knowledgebase^27^ (e.g. presence of a transmembrane domain); and (iv) Gene Ontology (GO) annotations.^28^ Following the method used in Pietzner et al^15^, we employed Boruta feature selection,^29^ a random-forest-based machine learning approach, to identify factors that were predictive of Spearman’s rho. We included a total of 88 features in Boruta (for GO, the top 10 most annotated terms from each GO category), with a p-value threshold of 0.01 and maximal number of 50,000 runs. An importance measure was generated for each variable after each run. For features confirmed by Boruta, we further tested their associations with Spearman’s rho using linear regression to determine the direction of their effect.

We performed GWAS of each protein, with genotyping array version, age, age^2^, sex, study area, and ten national genomic principal components included as covariates. GWAS were conducted using BOLT-LMM^30^ (SNPTEST^31^ when BOLT-LMM failed) for OLINK and REGENIE^32^ for SomaScan. Only SNPs with INFO>0.3 and minor allele count (MAC)>20 were included. Associated loci were defined by genome-wide significant variants (*p*<5×10^-8^) after linkage disequilibrium (LD) clumping using PLINK^33,34^ (initial window +/- 10 Mbp [40 Mbp for the MHC region], *p*<0.05, r^2^>0.05), with an internal LD reference of 40,000 unrelated CKB participants. Overlapping loci were merged and extended by +/- 10 kb, and the variant with the lowest p-value was identified as the sentinel variant. pQTLs within 500 kb on either side of the protein-encoding gene were defined as *cis-*pQTLs, while pQTLs outside this window were defined as *trans-*pQTLs. We performed fine-mapping using SuSiE (v0.12.16)^35^ to identify independent 95% credible sets. Colocalisation was performed using coloc (v5.2.1)^36^ for proteins with at least one *cis*-pQTL in either platform, to identify shared signals between OLINK and SomaScan with a posterior probability>0.8 across fine-mapped credible sets.

We conducted linear regression analyses to examine cross-sectional associations between protein levels and selected baseline traits, with scaled protein levels (i.e. divided by SD) as the outcome and traits of interest (e.g. BMI, heart rate, smoking) as explanatory variables. The models were adjusted for age, age^2^, sex, ambient temperature (at sample collection) and its square, time since last meal and its square, and plate ID. We employed the Benjamini–Hochberg method to control the false discovery rate (FDR) in multiple testing within each trait and each platform.^37^ The main analyses included all participants with additional sensitivity analyses restricted to the subcohort participants only.

We used the Prentice pseudo-partial likelihood Cox regression method for analyses of associations of levels of proteins with incident IHD (ICD-10 codes: I20, I22-I25).^38^ All models were adjusted for age, age^2^, sex, ambient temperature and its square, time since last meal and its square, plate ID, education, physical activity, alcohol intake, smoking, SBP, diabetes, and BMI. Sub-cohort participants were censored at time of any other IHD diagnosis.^39^

For risk prediction of IHD, we used LASSO logistic regression models using three sets of proteins: (i) all matched proteins; (ii) significant proteins in each platform after FDR correction; (iii) significant proteins that were shared between platforms. We calculated the Harrell’s concordance index (C-statistic)^40–42^ for each model using Prentice weighting^43^ and stratified the calculation by region. The C-statistic assesses a model’s discrimination ability within region and was estimated using repeated 10- fold cross-validation with 5 repeats in the present study. We used the net reclassification index (NRI)^44,45^ to assess the improvement gained by adding different sets of proteins to the conventional risk factors risk prediction model for IHD (including age, sex, smoking, type 2 diabetes, SBP, and waist circumference) previously developed in CKB.^46^ All analyses were conducted in R.^47^

## Results

Of the 3,976 participants in the study (one participant excluded due to insufficient sample volume), 53.7% were female and the mean (SD) age at baseline was 60.3 (11.5) years and the mean (SD) BMI was 23.9 (3.6) kg/m^2^. At sample collection, the mean ambient temperature was 15.7°C (SD 10.5), and the mean time since last meal was 4.7 (SD 4.7) hours. Further details of participant characteristics by case-subcohort ascertainment are shown in **Table 1**.

**Table 1:**
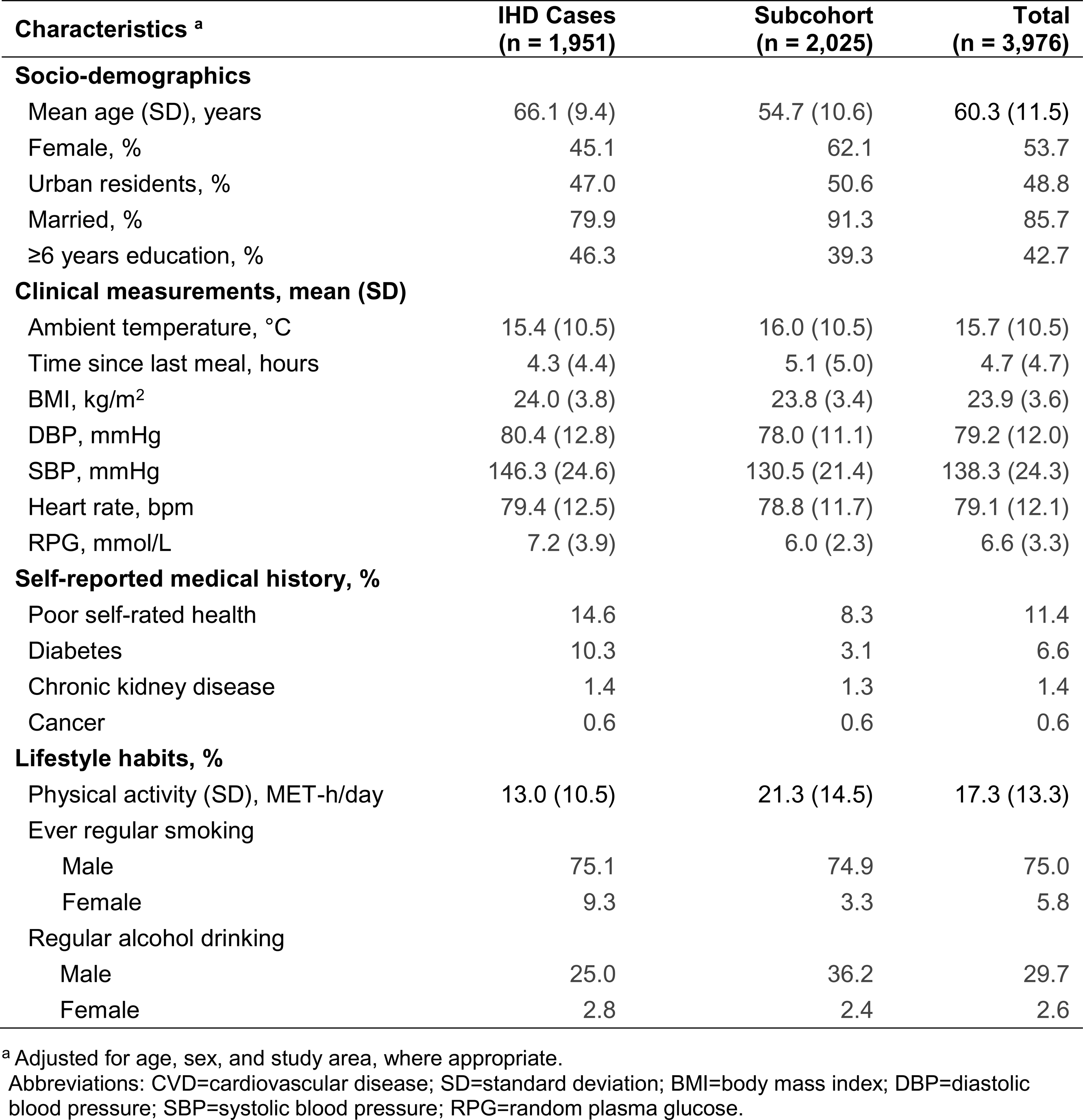
Baseline characteristics of IHD cases and subcohort participants who had no prior history of CVD at baseline.

| Characteristics <sup>a</sup> | IHD Cases<br>(n = 1,951) | Subcohort<br>(n = 2,025) | Total<br>(n = 3,976) |
| --- | --- | --- | --- |
| **Socio-demographics** |  |  |  |
| Mean age (SD), years | 66.1 (9.4) | 54.7 (10.6) | 60.3 (11.5) |
| Female, % | 45.1 | 62.1 | 53.7 |
| Urban residents, % | 47.0 | 50.6 | 48.8 |
| Married, % | 79.9 | 91.3 | 85.7 |
| ≥6 years education, % | 46.3 | 39.3 | 42.7 |
| **Clinical measurements, mean (SD)** |  |  |  |
| Ambient temperature, °C | 15.4 (10.5) | 16.0 (10.5) | 15.7 (10.5) |
| Time since last meal, hours | 4.3 (4.4) | 5.1 (5.0) | 4.7 (4.7) |
| BMI, kg/m<sup>2</sup> | 24.0 (3.8) | 23.8 (3.4) | 23.9 (3.6) |
| DBP, mmHg | 80.4 (12.8) | 78.0 (11.1) | 79.2 (12.0) |
| SBP, mmHg | 146.3 (24.6) | 130.5 (21.4) | 138.3 (24.3) |
| Heart rate, bpm | 79.4 (12.5) | 78.8 (11.7) | 79.1 (12.1) |
| RPG, mmol/L | 7.2 (3.9) | 6.0 (2.3) | 6.6 (3.3) |
| **Self-reported medical history, %** |  |  |  |
| Poor self-rated health | 14.6 | 8.3 | 11.4 |
| Diabetes | 10.3 | 3.1 | 6.6 |
| Chronic kidney disease | 1.4 | 1.3 | 1.4 |
| Cancer | 0.6 | 0.6 | 0.6 |
| **Lifestyle habits, %** |  |  |  |
| Physical activity (SD), MET-h/day | 13.0 (10.5) | 21.3 (14.5) | 17.3 (13.3) |
| Ever regular smoking |  |  |  |
| Male | 75.1 | 74.9 | 75.0 |
| Female | 9.3 | 3.3 | 5.8 |
| Regular alcohol drinking |  |  |  |
| Male | 25.0 | 36.2 | 29.7 |
| Female | 2.8 | 2.4 | 2.6 |

<sup>a</sup> Adjusted for age, sex, and study area, where appropriate.
Abbreviations: CVD=cardiovascular disease; SD=standard deviation; BMI=body mass index; DBP=diastolic blood pressure; SBP=systolic blood pressure; RPG=random plasma glucose.

After excluding duplicated reagents and reagents without corresponding UniProt IDs, there were 2,923 OLINK Explore 3072 reagents (mapped to 2,923 UniProt IDs), and 7,301 SomaScan Assay v4.1 SOMAmers targeting human proteins (mapped to 6,397 UniProt IDs). We identified 2,749 OLINK-SomaScan reagent pairs, corresponding to 2,168 UniProt IDs (**Figure 1**), including 1,694 reagent pairs for which one unique SOMAmer was matched to one unique OLINK reagent.

**Figure 1:**
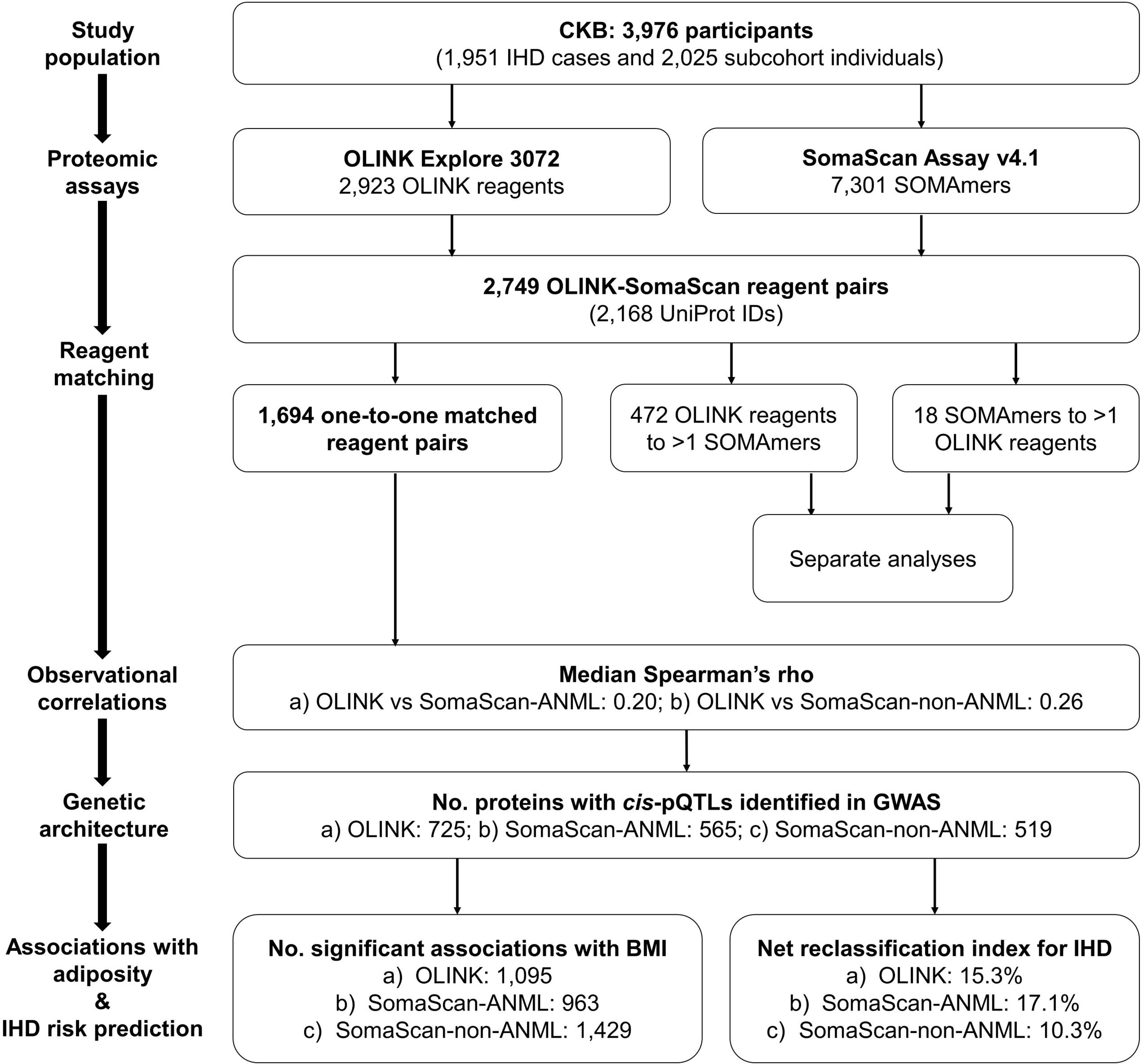
Summary of study design, analytic approaches and key findings. Main analyses were conducted on 1,694 one-to-one matched OLINK-SomaScan reagent pairs in 3,976 CKB participants. One subcohort participant was excluded due to insufficient sample volume. Results were corrected for multiple testing using false discovery rate within each platform. Risk prediction models for IHD were built using conventional risk factors (age, sex, smoking, T2D, SBP, and waist circumference) and significantly associated proteins. Abbreviations: CKB=China Kadoorie Biobank; ANML=adaptive normalization by maximum likelihood; BMI=body mass index; IHD=ischaemic heart disease.

### Observational correlations and associated factors

For the 1,694 one-to-one matched OLINK-SomaScan reagent pairs, we found a median rho of 0.20 between OLINK and SomaScan-ANML proteins, and a median rho of 0.26 between OLINK and SomaScan-non-ANML proteins. Histograms suggest a bimodal distribution of Spearman’s rho, with a peak centred around 0 and another closer to 0.8 (**Figure 2a**). There was a strong correlation between SomaScan-ANML and SomaScan-non-ANML data (median rho=0.84; **eFigure 1**). Analysis restricted to 2,025 subcohort participants yielded similar results (**eFigure 2**). Log-transformation of the SomaScan data increased the median Pearson’s r with OLINK from 0.05/0.08 to 0.15/0.20 (for ANML/non-ANML; **eFigure 3**). Therefore, all subsequent analyses were based on natural log-transformed SomaScan data.

**Figure 2:**
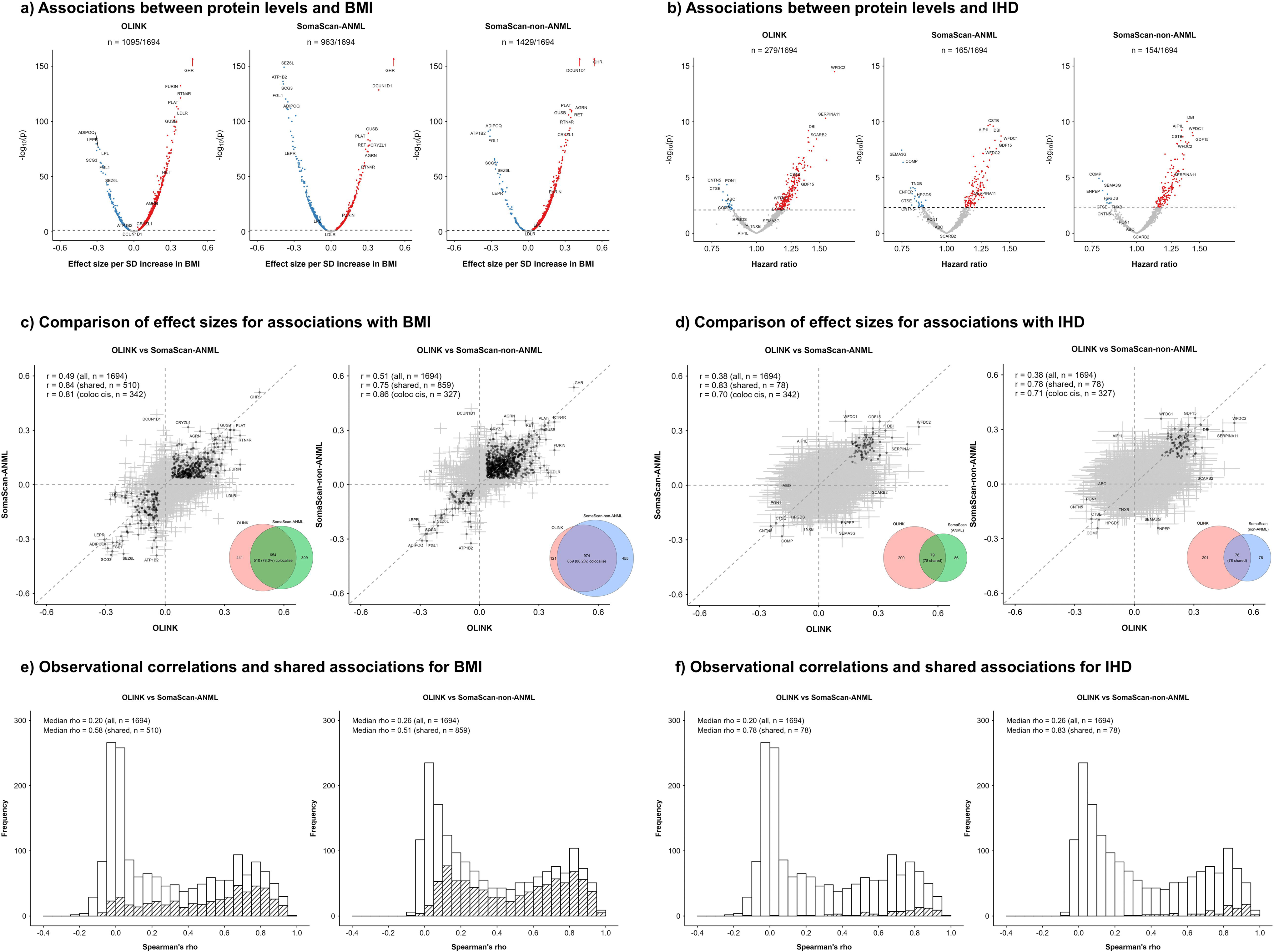
Observational correlations, associated factors, and comparison of pQTLs for 1,694 proteins measured using both OLINK and SomaScan platforms. a) Spearman’s correlation coefficients (rho) of protein levels between OLINK and SomaScan, with shaded areas indicating proteins with colocalising *cis*- pQTLs. b) Features predictive of Spearman’s rho and their importance in Boruta feature selection. Colours indicate the direction of their associations with Spearman’s rho. Two features were selected using non-ANML data but rejected using ANML data. c) Number of sentinel pQTLs identified in each platform, with shaded areas indicating the number of proteins with pQTLs. d) Number of proteins with *cis-*pQTLs identified in each platform and proteins with colocalising *cis*-pQTLs across platforms. Abbreviations: ANML=adaptive normalization by maximum likelihood; pQTL=protein quantitative trait loci.

**Figure 3:**
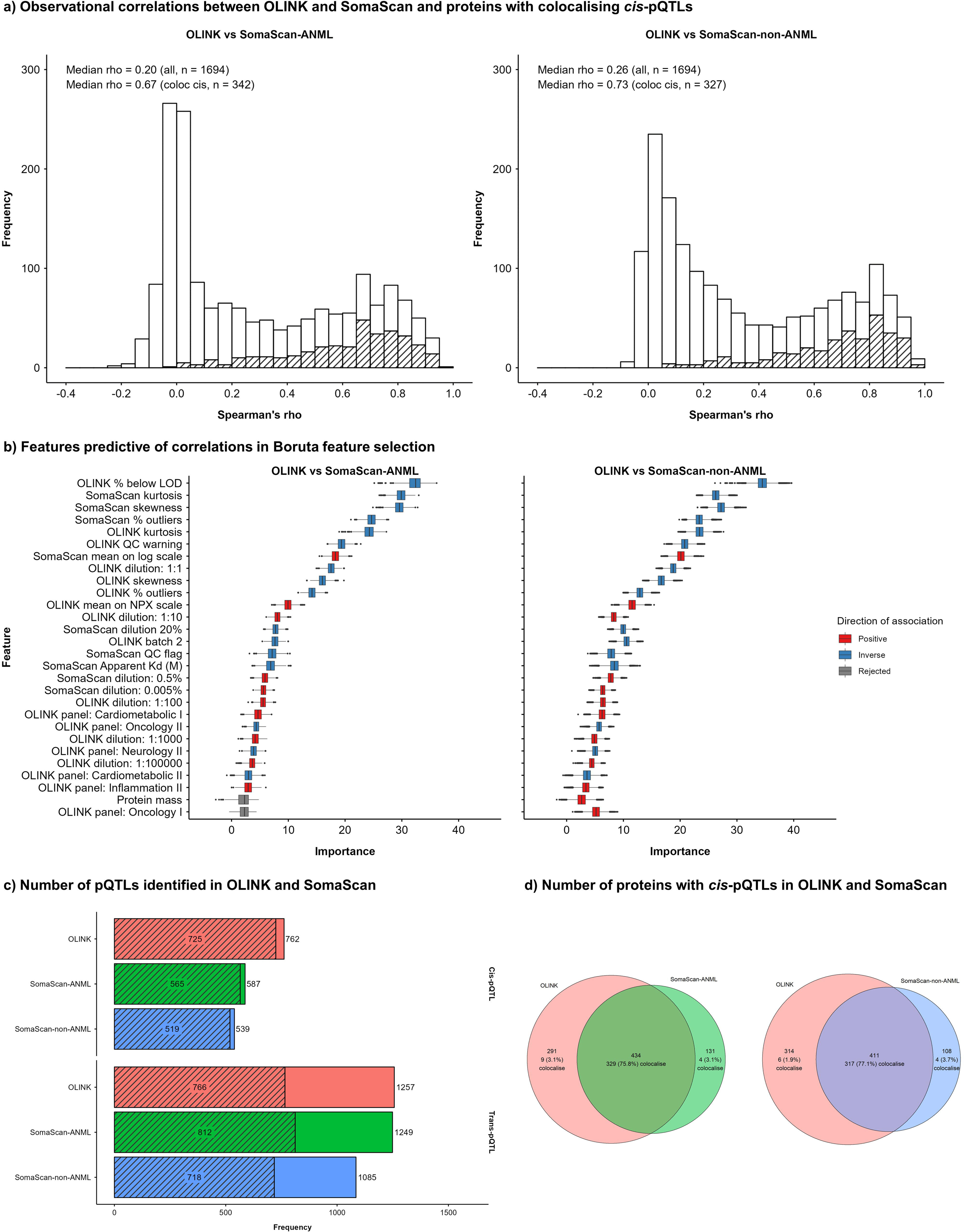
Comparison of number of proteins significantly associated with BMI and risk of incident IHD and their effect sizes between OLINK and SomaScan platforms. Associations between protein levels and BMI (a) and IHD (b).Comparison of effect sizes (beta coefficients) between OLINK and SomaScan for associations with BMI (c) and IHD (d). Spearman’s correlation coefficients of protein levels between OLINK and SomaScan, with shaded areas indicating shared associations for BMI (e) and IHD (f). Results were corrected for multiple testing using false discovery rate within each platform. Abbreviations: ANML=adaptive normalization by maximum likelihood; BMI=body mass index; IHD=ischaemic heart disease.

Using Boruta feature selection, several assay- and sample-related factors were predictive of Spearman’s correlation coefficients (**Figure 2b**). Higher protein abundance (i.e. higher mean values and dilution factors; **eFigures 4 and 5**) and higher data quality (i.e. lower % below LOD and lower % of samples with QC warnings) were predictive of higher correlations between platforms. Negative skewness (more values on the right side of the distribution) and platykurtic distributions (fewer extreme values, also indicated by lower % outliers) data were also associated with stronger correlations. Proteins in OLINK batch 1 (typically higher in abundance) were predictive of higher Spearman’s rho. The findings were consistent across ANML and non-ANML SomaScan data. In contrast, protein characteristics retrieved from the UniProt Knowledgebase and GO annotations were generally not predictive of correlations (**eTables 1 and 2**).

**Figure 4:**
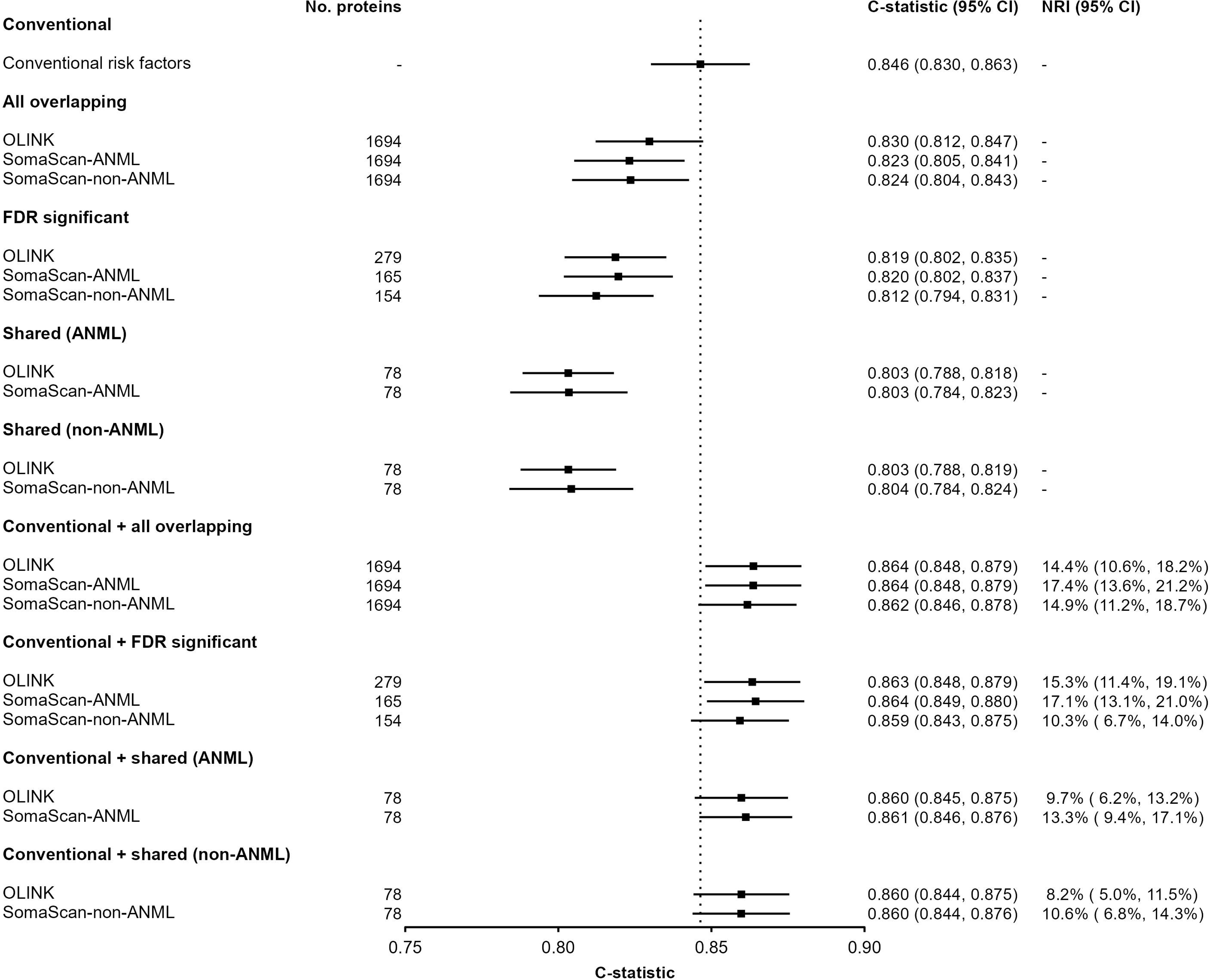
Performance of proteins measured using OLINK and SomaScan platforms for prediction of incident IHD. Conventional risk factors for cardiovascular disease included age, sex, smoking, T2D, SBP, and waist circumference. For each platform, three sets of proteins were used to construct risk prediction models: 1) all overlapping proteins; 2) out of the overlapping proteins, significant proteins after false discovery rate correction; 3) significant proteins that were shared between OLINK and SomaScan. Abbreviations: ANML=adaptive normalization by maximum likelihood; IHD=ischaemic heart disease; NRI=net reclassification index. FDR: false discovery rate.

### Comparisons of pQTLs

In GWAS of the 1,694 reagent pairs, there were 725 (42.8%), 565 (33.4%), and 519 (30.6%) proteins with *cis-*pQTLs identified in OLINK, SomaScan-ANML, and SomaScan-non-ANML, respectively, corresponding to 762, 587, and 539 sentinel *cis*-pQTL variants (**Figure 2c**). Moreover, there were 766 (45.2%), 812 (47.9%), and 718 (42.4%) proteins with *trans-*pQTLs, respectively, corresponding to 1,257, 1,249, and 1,085 sentinel *trans*-pQTL variants. The comparisons of −log10(p-value) for pQTL sentinel variants between platforms are shown in **eFigure6**.

In colocalisation analysis of 856 proteins with *cis*-pQTLs discovered in either OLINK or SomaScan-ANML (**Figure 2d**), 342 (40.0%) proteins had *cis-*pQTLs which colocalised between platforms. When restricting the analysis to 434 proteins with *cis-* pQTLs identified in *both* platforms, 329 (75.8%) showed evidence for colocalisation between OLINK and SomaScan-ANML platforms. In contrast, for proteins with *cis-* pQTLs identified in only one platform, only 3.1% showed evidence for colocalisation with the other platform. Proteins with colocalising *cis-*pQTLs also had higher between-platform correlations (median rho=0.67; **Figure 2a**). Colocalisation analysis of *cis*-pQTLs between OLINK and SomaScan-non-ANML yielded similar results (**Figure 2d**). The proportion of *cis*-pQTLs that colocalised was high between ANML and non-ANML SomaScan data (**eFigure 7**).

### Associations with adiposity and other traits

Overall BMI was significantly associated at FDR<0.05 with 1,095 (64.6%), 963 (56.8%), and 1,429 (84.4%) proteins, respectively, from OLINK, SomaScan-ANML, and SomaScan-non-ANML data (**Figure 3a**). Applying Bonferroni correction reduced the number of significant proteins proportionally across different platforms (**eFigure 8**). There was a moderate correlation of effect sizes (beta coefficients) for BMI between the platforms (Pearson’s r=0.49 [95%CI: 0.46 to 0.53] for ANML and 0.51 [95%CI: 0.47 to 0.54] for non-ANML with OLINK) (**Figure 3c**), and strongly correlated effect sizes (r=0.95; 95%CI: 0.95 to 0.95) between SomaScan-ANML and SomaScan-non-ANML data (**eFigure 9**).

Of the 654 BMI-associated proteins in both OLINK and SomaScan-ANML, 510 (78.0%) showed directionally consistent (i.e. shared) associations with BMI. For those with shared associations, the correlation of effect sizes between platforms was high (r=0.84; 95%CI: 0.82 to 0.87; **Figure 3c**). Proteins with shared associations with BMI had higher median Spearman’s correlations across platforms (**Figure 3e**). Among 342 proteins with colocalising *cis-*pQTLs between OLINK and SomaScan-ANML, there was a high between-platform correlation of effect sizes for BMI (r=0.81; 95%CI: 0.77 to 0.85), with 140 (40.9%) having shared associations with BMI. The results were similar for comparisons between OLINK and SomaScan-non-ANML platforms (**Figure 3**).

The associations of proteins with a range of other traits are shown in **eFigures 10-19**.

### Associations with incident IHD

Overall, 279 (16.4%), 165 (9.7%) and 154 (9.1%) proteins were significantly associated at FDR<0.05 with risk of IHD from OLINK, SomaScan-ANML and SomaScan-non-ANML data, respectively (**Figure 3b**). There were fewer significant associations with IHD after applying Bonferroni correction (**eFigure 20**). Of the 79 proteins that were significantly associated with IHD in both OLINK and SomaScan-ANML, 78 (98.7%) showed directionally concordant (shared) associations. For all overlapping proteins (n=1694), the between-platform correlation (Pearson’s r) of effect sizes was 0.38 (95%CI: 0.34 to 0.42), which increased to 0.83 (95%CI: 0.75 to 0.89) when restricted to proteins with shared associations with IHD (**Figure 3d**).

Proteins with shared associations with IHD had higher between-platform correlations (**Figure 3f**). When restricting the analyses to 342 proteins with colocalising *cis-* pQTLs, 40 (11.7%) had shared associations with IHD with a strong correlation of effect sizes (r=0.70; 95%CI: 0.64 to 0.75). The comparisons between OLINK and SomaScan-non-ANML platform were similar to those between OLINK and SomaScan-ANML (**Figure 3**), and the ANML and non-ANML SomaScan data had remarkably consistent effect sizes with BMI (r=0.98; 95%CI: 0.98 to 0.98; **eFigure 9**).

For prediction of incident IHD, models with more proteins typically performed better than those with fewer proteins, with the C-statistics ranging from 0.803 to 0.830 (**Figure 4**), compared with 0.846 (95%CI: 0.830 to 0.863) for the risk prediction model that only included conventional risk factors. The addition of proteins to the conventional model increased the C-statistics to about 0.860 irrespective of the platform or number of proteins included. Although the NRI did not differ notably between different platforms, there was a tendency for SomaScan-ANML proteins to have greater NRI values than OLINK proteins, even with fewer proteins considered (e.g. 17.1% vs 15.3% for 165 and 279 FDR-significant proteins in SomaScan-ANML and OLINK, respectively).

### Non-one-to-one matched reagent pairs

Of the remaining 474 proteins matched to more than one reagent, 472 were targeted by one OLINK reagent and by 2 to 9 SOMAmers (1,037 unique SOMAmers in total), resulting in 1,053 OLINK-SomaScan reagent pairs. We found a moderate correlation between protein levels measured by different SOMAmers targeting the same protein (median rho=0.18/0.46 for ANML/non-ANML; **eFigure 21**). Likewise, there was only a moderate correlation between OLINK and SomaScan reagent pairs (median rho=0.31/0.37 for ANML/non-ANML; **eFigure 22**). In GWAS of these 474 proteins, *cis*-pQTLs were identified for 258 OLINK reagents, 427 ANML-SOMAmers, and 400 non-ANML-SOMAmers, and *trans*-pQTLs were identified for 257, 566, and 500 reagents, respectively. We found evidence for colocalisation of *cis*-pQTLs for 181/174 proteins between OLINK reagents and varying numbers of their matched ANML- and non-ANML-SOMAmers (**eTables 3 and 4**).

A further 18 proteins were targeted by one SOMAmer and two OLINK reagents (22 unique OLINK reagents in total), resulting in 36 OLINK-SomaScan reagent pairs. There were moderate correlations between OLINK reagents targeting the same protein (median r=0.31; **eFigure 23**) and between OLINK and SomaScan reagent pairs (median rho=0.07/0.16 for ANML/non-ANML; **eFigure 24**). In GWAS of these 18 proteins, *cis*-pQTLs were identified by 8 OLINK reagents, 5 ANML-SOMAmers, and 6 non-ANML-SOMAmers, while *trans*-pQTLs were identified by 15, 9, and 6 reagents, respectively. There was evidence for colocalisation of *cis*-pQTLs for only two SOMAmers with one of their two matched OLINK reagents.

## Discussion

In this large-scale direct comparative analysis of 2,168 proteins assayed by both OLINK and SomaScan platforms in Chinese adults, we found only modest correlations between plasma levels of proteins measured by each platform. In GWAS, OLINK-proteins were more likely to have *cis-*pQTLs compared with SomaScan-proteins, with >70% proteins with overlapping *cis*-pQTLs colocalised. Both platforms yielded comparable numbers of proteins significantly associated with adiposity. There were more OLINK than SomaScan proteins significantly associated with incident IHD, but their performance for risk prediction of IHD was similar, with SomaScan proteins, irrespective of the number of proteins considered, showing slightly higher NRI values for IHD when added to conventional IHD risk factors.

Previous studies designed to compare OLINK and SomaScan platforms had varying numbers of participants (10∼1,514) and numbers of overlapping proteins (425∼1,848). However, most studies reported only moderate correlations (median rho≈0.4) of protein levels between each platform.^13–16^ Two previous studies examined the concordance of pQTLs identified by the two platforms in the same sample, with one reporting half of the proteins with *cis*-pQTLs having signals discovered in both,^16^ while another reported that 64% of genomic region-protein associations were shared based on LD.^15^ The recent study by Eldjarn et al. reported that about half of the *cis*-pQTL signals in one platform had a corresponding pQTL in the other, although their pQTLs were identified in two different datasets.^17^ In contrast with the present study, previous studies did not analyse proteins separately targeted by multiple reagents, which showed different patterns of associations, nor assessed the consistency of pQTLs obtained by colocalisation analyses. In the present study, we found more modest correlations (median rho=0.20) between the two platforms than previous studies, possibly reflecting the inclusion of a higher proportion of low-abundance proteins in the present study. Indeed, in analyses stratified by protein abundance, there were stronger correlations (median rho≈0.6) for more highly abundant proteins (i.e. those with dilution factor<10%). In the present colocalisation analyses, we found that about 40% of the proteins with *cis*-pQTLs obtained by *either* platform colocalised between OLINK and SomaScan, with the proportion increasing to 70% when analyses were restricted to those with *cis*-pQTLs discovered in *both* platforms. In this East Asian population, we were also able to identify pQTLs that had not been previously reported in other populations due to MAF differences. For example, the *cis*-pQTL sentinel variant (rs671) for ALDH2 has an MAF=0.20 in East Asians but <0.001 in other populations, and is known to have a significant impact on alcohol metabolism.^48,49^ The missense variant rs76863441 (MAF=0.06 in East Asians but <0.001 in other populations) was a *cis*-pQTL for PLA2G7, which was found to be related to inflammation, CVD, and life span.^50,51^ The two *cis*-pQTLs were found in both OLINK and SomaScan and colocalised between platforms (**eFigures 25 and 26**), which should facilitate future research on such questions.

Few previous studies have systematically compared the associations of OLINK and SomaScan proteins with traits and risks of disease outcomes. In the present study, the two platforms yielded comparable numbers of significant associations with BMI and other traits. For incident IHD, however, the OLINK platform yielded more proteins that were significantly associated with disease risk than SomaScan. Nevertheless, most (∼80%) proteins that were significantly associated with IHD in both platforms showed directionally consistent associations, with a high (r≥0.75) correlation of effect sizes. We found that proteins with high correlations between platforms or with colocalising *cis*-pQTLs tended to produce reproducible associations with traits and disease risk across platforms, as illustrated by *GHR* (associated with BMI in both platforms) and *GDF15* (associated with IHD in both platforms). Both proteins showed high concordance in their observational correlations and *cis*-pQTLs, and their associations with adiposity or with CVD have been previously reported.^52–54^

The present study was the first study to directly compare the performance of OLINK and SomaScan proteins for risk prediction of major diseases in the same population. Although proteomic prediction models alone did not outperform the performance of the model with conventional risk factors for IHD in analyses of a set of proteins captured by both platforms, the addition of proteins, irrespective of the number considered, to conventional risk factors significantly improved risk prediction of IHD. Moreover, the NRI for SomaScan proteins, especially SomaScan-ANML, were somewhat higher than those obtained for OLINK proteins, which persisted even with a smaller number of FDR-significant proteins (165 vs 279 proteins; NRI: 17.1% vs 15.3%). Importantly, the present analyses focused on the 1,694 overlapping proteins and the SomaScan platform captures more proteins than OLINK platform, which may yield improved risk prediction of IHD (and other diseases) when all proteins are considered.^55,56^

In contrast with the present study, previous studies have focused on comparisons between OLINK and SomaScan-ANML, without due consideration of SomaScan-non-ANML. Although most results were highly consistent, we found that SomaScan-non-ANML yielded a larger number of significant associations with BMI (1,429 vs 963) and particular baseline traits. The ANML procedure standardises the overall signal of each sample to an external reference, and while improving the data quality, this process may also suppress true extreme signals or outliers that are present in the general population, resulting in fewer significant associations.^7,8^ For incident IHD, however, the findings differed, with comparable numbers of significant associations for SomaScan-ANML and SomaScan-non-ANML, but slightly greater NRI detected by SomaScan-ANML for risk prediction. These findings warrant corroboration in other ancestry populations given their potential importance for population health research.

Consistent with previous studies,^15^ our Boruta feature selection analyses demonstrated that protein abundance and data quality (e.g. % below LOD, % outliers, QC warning) were the two most important factors that accounted for the concordance between measurements as well as differences in downstream genetic and observational analyses between OLINK and SomaScan platforms. Conversely, protein characteristics based on annotations from UniProt Knowledgebase and Gene Ontology contributed little to the differences between platforms, at least for the 1,694 proteins with one-to-one matched single reagents. Nevertheless, differences in protein measurements may still be influenced by reagents binding to different proteoforms of the same protein, leading to epitope effects and discordant findings in pQTL analysis (i.e. genetic variation influencing reagent binding to protein epitopes due to changes in protein structure).^10,17^ Further in-depth analyses of 474 proteins matched to more than one reagent could explore possible epitope effects. For example, not all SOMAmers targeting TNC showed evidence for colocalisation of *cis*-pQTLs with the corresponding OLINK reagent (**eFigures 27 and 28**). This could be a result of those reagents targeting different structures of TNC, a protein known to have different isoforms.^57,58^ In the absence of any data on proteoforms for either platform, it was not possible to reliably assess this hypothesis.

The present study is the largest study to date that compares the diagnostic utility of antibody and aptamer-based proteomic platforms in identical blood samples. We assessed the agreement between platforms using both observational and genetic analyses, and compared analytical performance for their associations with traits and IHD, in addition to utility for risk prediction of IHD. In addition, this is the first such study to be conducted in East Asians, and this has enabled the identification of pQTLs that were not previously reported in other ancestry populations. However, the present study also had several limitations. First, we did not compare coefficients of variation as a measure of accuracy, as these are strongly influenced by the data distribution. Moreover, we were unable to adopt the CV ratio (i.e. CV in repeated samples divided by CV in unrelated samples) as proposed by Eldjarn et al to compare accuracy between platforms,^17^ due to the small number of duplicates of proteins/samples measured in the study. Second, the analyses chiefly focused on one-to-one matched OLINK-SomaScan reagent pairs, and further analyses of proteins assayed using different reagents targeting the same proteins may still offer additional insights, particularly analysis of possible epitope effects.^15^ Third, we only compared 2,168 overlapping proteins matched to the same UniProt ID based on the previous version of each platform. Recently, the number of proteins measured by OLINK has increased to over 5,000, while the latest SomaScan platform includes about 11,000 aptamers, and this is likely to increase the number of overlapping proteins between the platforms. Finally, we only compared two major affinity-based platforms. Future studies will be required to compare agreement with measurements from mass spectrometry platforms, which remains the ‘gold standard’ for protein identification and quantification. With advances in mass spectrometry that increase assay throughput,^59,60^ future comparative studies of mass spectrometry with affinity-based platform(s) are now warranted.

Overall, this study assessed the analytical performance of two affinity-based proteomic platforms in a large study of Chinese adults. Each platform had different strengths and limitations and both were complementary with each other. The selection of platforms in future studies may depend on the study purpose (e.g. mechanistic investigation vs risk prediction), analytical approach (e.g. observational vs genetic), preferred breadth of coverage of the platform, and overall cost-effectiveness. As affinity-based and mass spectrometry technologies are still growing and will likely capture more overlapping proteins between platforms, further studies are still needed to compare, both directly and indirectly, the utilities of different platforms that will inform large-scale population and clinical research.

## Supporting information

eTables and eFigures

## Data Availability

The China Kadoorie Biobank (CKB) is a global resource for the investigation of lifestyle, environmental, blood biochemical and genetic factors as determinants of common diseases. The CKB study group is committed to making the cohort data available to the s cientific community in China, the UK and worldwide to advance knowledge about the causes, prevention and treatment of disease. Full details of what data is currently available to open access users and how to apply for it, visit: http://www.ckbiobank.org/site/Data+Access. Researchers who are interested in obtaining the raw data from the China Kadoorie Biobank study that underlines this paper should contact. A research proposal will be requested to ensure that any analysis is performed by bona fide researchers and - where data is not currently available to open access researchers - is restricted to the topic covered in this paper.

## Acknowledgments

The chief acknowledgment is to the participants, the project staff, and the China CDC and its regional offices for assisting with the fieldwork. We thank Judith Mackay in Hong Kong; Yu Wang, Gonghuan Yang, Zhengfu Qiang, Lin Feng, Maigeng Zhou, Wenhua Zhao, and Yan Zhang in China CDC; Lingzhi Kong, Xiucheng Yu, and Kun Li in the Chinese Ministry of Health for assisting with conduct and organization of the study.

## Ethics approval

The China Kadoorie Biobank (CKB) complies with all required ethical standards for medical research on human subjects. Ethical approvals were obtained and been maintained by the relevant institutional ethical research committees in the UK and China.

## Consent to participate/publication

All participants provided written informed consent.

## Data sharing statement

The China Kadoorie Biobank (CKB) is a global resource for the investigation of lifestyle, environmental, blood biochemical and genetic factors as determinants of common diseases. CKB is committed to making the cohort data available to the scientific community and for information on data currently available to open access users and how to apply for it, please visit: http://www.ckbiobank.org/site/Data+Access.

## Code availability

Custom code was used for statistical analyses will be made available to external researchers upon reasonable request to bona fide researchers.

## Author contributions

BW, RW, RC, IM, and ZC contributed to the concept and design of this study. BW, AP, MM, NW conducted the statistical analyses and BW, IM and ZC drafted the manuscript. BW, AP, MM, NW, PY, SS, AI, CK, HF, KL, YC, HD, DA, DVS, CY, DS, JL, MH, LL, DB, RCo, RW, RC, IM, and ZC were involved in the planning, acquisition and interpretation of data. HF, KL, YC, HD, DA, and DS and provided administrative, technical, or material support. All authors provided critical revision of the manuscript for important intellectual content. BW, AP, MM, NW, IM and ZC are the guarantors of this work and take responsibility for the integrity and accuracy of the data analysis. IM and ZC supervised the work.

## Abbreviations

ALDH2: Aldehyde Dehydrogenase 2 Family Member
ANML: Adaptive normalization by maximum likelihood
BMI: Body mass index
CKB: China Kadoorie Biobank
CVD: Cardiovascular disease
DBP: Diastolic blood pressure
FDR: False discovery rate
GDF15: Growth Differentiation Factor 15
GHR: Growth Hormone Receptor
GO: Gene Ontology
GWAS: Genome-wide association study
HDL: High-density lipoprotein
IHD: Ischaemic heart disease
LD: Linkage disequilibrium
LOD: Limit of detection
MAC: Minor allele count
MAF: Minor allele frequency
MHC: Major histocompatibility complex
NRI: Net Reclassification Index
NPX: Normalized Protein eXpression
NRI: Net reclassification index
PLA2G7: Phospholipase A2 Group VII
pQTL: Protein quantitative trait loci
QC: Quality control
RFU: Relative fluorescence units
ROC: Receiver operating characteristic
SBP: Systolic blood pressure
SOMAmer: Slow offrate modified aptamers
TNC: Tenascin C

## References

1. Domon B, Aebersold R. Mass spectrometry and protein analysis. Science (1979). 2006;312(5771):212-217. doi:10.1126/SCIENCE.1124619

2. Keshishian H, Burgess MW, Specht H, et al. Quantitative, multiplexed workflow for deep analysis of human blood plasma and biomarker discovery by mass spectrometry. Nature Protocols 2017 12:8. 2017;12(8):1683–1701. doi:10.1038/nprot.2017.054

3. Tognetti M, Sklodowski K, Müller S, et al. Biomarker Candidates for Tumors Identified from Deep-Profiled Plasma Stem Predominantly from the Low Abundant Area. J Proteome Res. 2022;21(7):1718–1735. doi:10.1021/ACS.JPROTEOME.2C00122

4. Suhre K, McCarthy MI, Schwenk JM. Genetics meets proteomics: perspectives for large population-based studies. Nature Reviews Genetics 2021 22:1. 2021;22(1):19–37. doi:10.1038/s41576-020-0268-2

5. Assarsson E, Lundberg M, Holmquist G, et al. Homogenous 96-Plex PEA Immunoassay Exhibiting High Sensitivity, Specificity, and Excellent Scalability. PLoS One. 2014;9(4):e95192. doi:10.1371/JOURNAL.PONE.0095192

6. Gold L, Ayers D, Bertino J, et al. Aptamer-Based Multiplexed Proteomic Technology for Biomarker Discovery. PLoS One. 2010;5(12):e15004. doi:10.1371/JOURNAL.PONE.0015004

7. Candia J, Daya GN, Tanaka T, Ferrucci L, Walker KA. Assessment of variability in the plasma 7k SomaScan proteomics assay. Scientific Reports 2022 12:1. 2022;12(1):1–12. doi:10.1038/s41598-022-22116-0

8. Candia J, Cheung F, Kotliarov Y, et al. Assessment of Variability in the SOMAscan Assay. Scientific Reports 2017 7:1. 2017;7(1):1–13. doi:10.1038/s41598-017-14755-5

9. Sun BB, Chiou J, Traylor M, et al. Genetic regulation of the human plasma proteome in 54,306 UK Biobank participants. bioRxiv. 2022;20:2022.06.17.496443. doi:10.1101/2022.06.17.496443

10. Ferkingstad E, Sulem P, Atlason BA, et al. Large-scale integration of the plasma proteome with genetics and disease. Nature Genetics 2021 53:12. 2021;53(12):1712–1721. doi:10.1038/s41588-021-00978-w

11. Koprulu M, Carrasco-Zanini J, Wheeler E, et al. Proteogenomic links to human metabolic diseases. Nature Metabolism 2023 5:3. 2023;5(3):516–528. doi:10.1038/s42255-023-00753-7

12. Xu Y, Ritchie SC, Liang Y, et al. An atlas of genetic scores to predict multi-omic traits. Nature 2023 616:7955. 2023;616(7955):123–131. doi:10.1038/s41586-023-05844-9

13. Haslam DE, Li J, Dillon ST, et al. Stability and reproducibility of proteomic profiles in epidemiological studies: comparing the Olink and SOMAscan platforms. Proteomics. 2022;22(13-14). doi:10.1002/PMIC.202100170

14. Raffield LM, Dang H, Pratte KA, et al. Comparison of Proteomic Assessment Methods in Multiple Cohort Studies. Proteomics. 2020;20(12):e1900278. doi:10.1002/PMIC.201900278

15. Pietzner M, Wheeler E, Carrasco-Zanini J, et al. Synergistic insights into human health from aptamer- and antibody-based proteomic profiling. Nature Communications 2021 12:1. 2021;12(1):1–13. doi:10.1038/s41467-021-27164-0

16. Katz DH, Robbins JM, Deng S, et al. Proteomic profiling platforms head to head: Leveraging genetics and clinical traits to compare aptamer- and antibody-based methods. Sci Adv. 2022;8(33). doi:10.1126/SCIADV.ABM5164

17. Eldjarn GH, Ferkingstad E, Lund SH, et al. Large-scale plasma proteomics comparisons through genetics and disease associations. Nature. Published online October 4, 2023. doi:10.1038/s41586-023-06563-x

18. Chen Z, Chen J, Collins R, et al. China Kadoorie Biobank of 0.5 million people: survey methods, baseline characteristics and long-term follow-up. Int J Epidemiol. 2011;40(6):1652–1666.

19. Yao P, Iona A, Kartsonaki C, et al. Conventional and genetic associations of adiposity with 1463 proteins in relatively lean Chinese adults. Eur J Epidemiol. Published online September 7, 2023. doi:10.1007/S10654-023-01038-9

20. Zhao H, Sun Z, Wang J, Huang H, Kocher JP, Wang L. CrossMap: a versatile tool for coordinate conversion between genome assemblies. Bioinformatics. 2014;30(7):1006–1007. doi:10.1093/BIOINFORMATICS/BTT730

21. Delaneau O, Marchini J, Zagury JF. A linear complexity phasing method for thousands of genomes. Nat Methods. 2012;9(2):179–181. doi:10.1038/nmeth.1785

22. Das S, Forer L, Schönherr S, et al. Next-generation genotype imputation service and methods. Nat Genet. 2016;48(10):1284–1287. doi:10.1038/ng.3656

23. Cong PK, Bai WY, Li JC, et al. Genomic analyses of 10,376 individuals in the Westlake BioBank for Chinese (WBBC) pilot project. Nature Communications 2022 13:1. 2022;13(1):1–15. doi:10.1038/s41467-022-30526-x

24. Walters RG, Millwood IY, Lin K, et al. Genotyping and population characteristics of the China Kadoorie Biobank. Cell Genomics. 2023;0(0):100361. doi:10.1016/J.XGEN.2023.100361

25. Olink. Data Normalization and Standardization.; 2021. Accessed November 7, 2023. https://olink.com/application/data-normalization-and-standardization/

26. SomaLogic. SomaScan® v4.0 and v4.1 Data Standardization.; 2021. Accessed November 7, 2023. https://somalogic.com/tech-notes/

27. Bateman A, Martin MJ, Orchard S, et al. UniProt: the Universal Protein Knowledgebase in 2023. Nucleic Acids Res. 2023;51(D1):D523–D531. doi:10.1093/NAR/GKAC1052

28. Carbon S, Douglass E, Dunn N, et al. The Gene Ontology Resource: 20 years and still GOing strong. Nucleic Acids Res. 2019;47(D1):D330–D338. doi:10.1093/NAR/GKY1055

29. Kursa MB, Rudnicki WR. Feature Selection with the Boruta Package. J Stat Softw. 2010;36(11):1–13. doi:10.18637/JSS.V036.I11

30. Loh PR, Tucker G, Bulik-Sullivan BK, et al. Efficient Bayesian mixed-model analysis increases association power in large cohorts. Nature Genetics 2015 47:3. 2015;47(3):284–290. doi:10.1038/ng.3190

31. Marchini J, Howie B, Myers S, McVean G, Donnelly P. A new multipoint method for genome-wide association studies by imputation of genotypes. Nature Genetics 2007 39:7. 2007;39(7):906–913. doi:10.1038/ng2088

32. Mbatchou J, Barnard L, Backman J, et al. Computationally efficient whole-genome regression for quantitative and binary traits. Nature Genetics 2021 53:7. 2021;53(7):1097–1103. doi:10.1038/s41588-021-00870-7

33. Chang CC, Chow CC, Tellier LC, Vattikuti S, Purcell SM, Lee JJ. Second-generation PLINK: rising to the challenge of larger and richer datasets. Gigascience. 2015;4:7. doi:10.1186/s13742-015-0047-8

34. Purcell SM, Neale B, Todd-Brown K, et al. PLINK: a tool set for whole-genome association and population-based linkage analyses. Am J Hum Genet. 2007;81(3):559–575. doi:10.1086/519795

35. Wang G, Sarkar A, Carbonetto P, Stephens M. A Simple New Approach to Variable Selection in Regression, with Application to Genetic Fine Mapping. J R Stat Soc Series B Stat Methodol. 2020;82(5):1273–1300. doi:10.1111/RSSB.12388

36. Giambartolomei C, Vukcevic D, Schadt EE, et al. Bayesian Test for Colocalisation between Pairs of Genetic Association Studies Using Summary Statistics. Williams SM, ed. PLoS Genet. 2014;10(5):e1004383. doi:10.1371/journal.pgen.1004383

37. Benjamini Y, Hochberg Y. Controlling the false discovery rate: a practical and powerful approach to multiple testing. Journal of the Royal statistical society: series B (Methodological). 1995;57(1):289–300.

38. Prentice RL. A Case-Cohort Design for Epidemiologic Cohort Studies and Disease Prevention Trials. Biometrika. 1986;73(1):1. doi:10.2307/2336266

39. Mazidi M, Wright N, Yao P, et al. Plasma Proteomics to Identify Drug Targets for Ischemic Heart Disease. J Am Coll Cardiol. 2023;82(20):1906–1920. doi:10.1016/J.JACC.2023.09.804/SUPPL_FILE/MMC1.DOCX

40. Harrell FE, Califf RM, Pryor DB, Lee KL, Rosati RA. Evaluating the Yield of Medical Tests. JAMA. 1982;247(18):2543–2546. doi:10.1001/JAMA.1982.03320430047030

41. Harrell FE, Lee KL, Califf RM, Pryor DB, Rosati RA. Regression modelling strategies for improved prognostic prediction. Stat Med. 1984;3(2):143–152. doi:10.1002/SIM.4780030207

42. Harrell FE, Lee KL, Mark DB. Multivariable prognostic models: issues in developing models, evaluating assumptions and adequacy, and measuring and reducing errors. Stat Med. 1996;15(4):361–387.

43. Sanderson J, Thompson SG, White IR, Aspelund T, Pennells L. Derivation and assessment of risk prediction models using case-cohort data. BMC Med Res Methodol. 2013;13(1):1–11. doi:10.1186/1471-2288-13-113/FIGURES/4

44. Pencina MJ, D’Agostino RB, D’Agostino RB, Vasan RS. Evaluating the added predictive ability of a new marker: from area under the ROC curve to reclassification and beyond. Stat Med. 2008;27(2):157–172. doi:10.1002/SIM.2929

45. McKearnan SB, Wolfson J, Vock DM, Vazquez-Benitez G, O’Connor PJ. Performance of the Net Reclassification Improvement for Nonnested Models and a Novel Percentile-Based Alternative. Am J Epidemiol. 2018;187(6):1327. doi:10.1093/AJE/KWX374

46. Yang S, Han Y, Yu C, et al. Development of a Model to Predict 10-Year Risk of Ischemic and Hemorrhagic Stroke and Ischemic Heart Disease Using the China Kadoorie Biobank. Neurology. 2022;98(23):e2307–e2317. doi:10.1212/WNL.0000000000200139

47. R Core Team. R: A Language and Environment for Statistical Computing. Published online 2022. http://www.r-project.org/index.html

48. Im PK, Wright N, Yang L, et al. Alcohol consumption and risks of more than 200 diseases in Chinese men. Nature Medicine 2023 29:6. 2023;29(6):1476–1486. doi:10.1038/s41591-023-02383-8

49. Millwood IY, Walters RG, Mei XW, et al. Conventional and genetic evidence on alcohol and vascular disease aetiology: a prospective study of 500 000 men and women in China. The Lancet. 2019;393(10183):1831–1842. doi:10.1016/S0140-6736(18)31772-0

50. Spadaro O, Youm Y, Shchukina I, et al. Caloric restriction in humans reveals immunometabolic regulators of health span. Science (1979). 2022;375(6581):671–677. doi:10.1126/SCIENCE.ABG7292/SUPPL_FILE/SCIENCE.ABG7292_MDAR_REPRODUCIBILITY_CHECKLIST.PDF

51. Huang F, Wang K, Shen J. Lipoprotein-associated phospholipase A2: The story continues. Med Res Rev. 2020;40(1):79–134. doi:10.1002/MED.21597

52. Wang D, Day EA, Townsend LK, Djordjevic D, Jørgensen SB, Steinberg GR. GDF15: emerging biology and therapeutic applications for obesity and cardiometabolic disease. Nature Reviews Endocrinology 2021 17:10. 2021;17(10):592–607. doi:10.1038/s41574-021-00529-7

53. Adela R, Banerjee SK. GDF-15 as a target and biomarker for diabetes and cardiovascular diseases: A translational prospective. J Diabetes Res. 2015;2015. doi:10.1155/2015/490842

54. Kopchick JJ, Berryman DE, Puri V, Lee KY, Jorgensen JOL. The effects of growth hormone on adipose tissue: old observations, new mechanisms. Nat Rev Endocrinol. 2020;16(3):135–146. doi:10.1038/S41574-019-0280-9

55. Hoogeveen RM, Pereira JPB, Nurmohamed NS, et al. Improved cardiovascular risk prediction using targeted plasma proteomics in primary prevention. Eur Heart J. 2020;41(41):3998–4007. doi:10.1093/EURHEARTJ/EHAA648

56. Nurmohamed NS, Belo Pereira JP, Hoogeveen RM, et al. Targeted proteomics improves cardiovascular risk prediction in secondary prevention. Eur Heart J. 2022;43(16):1569–1577. doi:10.1093/EURHEARTJ/EHAC055

57. Giblin SP, Midwood KS. Tenascin-C: Form versus function. Cell Adh Migr. 2015;9(1-2):48. doi:10.4161/19336918.2014.987587

58. Midwood KS, Hussenet T, Langlois B, Orend G. Advances in tenascin-C biology. Cellular and Molecular Life Sciences 2011 68:19. 2011;68(19):3175–3199. doi:10.1007/S00018-011-0783-6

59. Cui M, Cheng C, Zhang L. High-throughput proteomics: a methodological mini-review. Laboratory Investigation 2022 102:11. 2022;102(11):1170–1181. doi:10.1038/s41374-022-00830-7

60. Bian Y, Zheng R, Bayer FP, et al. Robust, reproducible and quantitative analysis of thousands of proteomes by micro-flow LC–MS/MS. Nature Communications 2020 11:1. 2020;11(1):1–12. doi:10.1038/s41467-019-13973-x

