## Supplementary material for "Comparative studies of genetic and phenotypic associations for 2,168 plasma proteins measured by two affinity-based platforms in 4,000 Chinese adults": eTables and eFigures

**Members of CKB Collaborative Group**

**International Steering Committee:** Junshi Chen, Zhengming Chen (PI), Robert Clarke, Rory Collins, Liming Li (PI), Chen Wang, Jun Lv, Richard Peto, Robin Walters.

**International Co-ordinating Centre, Oxford:** Daniel Avery, Maxim Barnard, Derrick Bennett, Ruth Boxall, Ka Hung Chan, Yiping Chen, Zhengming Chen, Jonathan Clarke, Robert Clarke, Huaidong Du, Ahmed Edris Mohamed, Hannah Fry, Simon Gilbert, Pek Kei Im, Andri Iona, Maria Kakkoura, Christiana Kartsonaki, Hubert Lam, Kuang Lin, James Liu, Mohsen Mazidi, Iona Millwood, Sam Morris, Qunhua Nie, Alfred Pozarickij, Paul Ryder, Saredo Said, Dan Schmidt, Becky Stevens, Iain Turnbull, Robin Walters, Baihan Wang, Lin Wang, Neil Wright, Ling Yang, Xiaoming Yang, Pang Yao.

**National Co-ordinating Centre, Beijing:** Xiao Han, Can Hou, Qingmei Xia, Chao Liu, Jun Lv, Pei Pei, Dianjanyi Sun, Canqing Yu

**10 Regional Co-ordinating Centres:**

**Guangxi** Provincial CDC: Naying Chen, Duo Liu, Zhenzhu Tang. Liuzhou CDC: Ningyu Chen, Qilian Jiang, Jian Lan, Mingqiang Li, Yun Liu, Fanwen Meng, Jinhuai Meng, Rong Pan, Yulu Qin, Ping Wang, Sisi Wang, Liuping Wei, Liyuan Zhou. **Gansu** Provincial CDC: Caixia Dong, Pengfei Ge, Xiaolan Ren. Maiji CDC: Zhongxiao Li, Enke Mao, Tao Wang, Hui Zhang, Xi Zhang. **Hainan** Provincial CDC: Jinyan Chen, Ximin Hu, Xiaohuan Wang. Meilan CDC: Zhendong Guo, Huimei Li, Yilei Li, Min Weng, Shukuan Wu. **Heilongjiang** Provincial CDC: Shichun Yan, Mingyuan Zou, Xue Zhou. Nangang CDC: Ziyan Guo, Quan Kang, Yanjie Li, Bo Yu, Qinai Xu. **Henan** Provincial CDC: Liang Chang, Lei Fan, Shixian Feng, Ding Zhang, Gang Zhou. Huixian CDC: Yulian Gao, Tianyou He, Pan He, Chen Hu, Huarong Sun, Xukui Zhang. **Hunan** Provincial CDC: Biyun Chen, Zhongxi Fu, Yuelong Huang, Huilin Liu, Qiaohua Xu, Li Yin. Liuyang CDC: Huajun Long, Xin Xu, Hao Zhang, Libo Zhang. **Jiangsu** Provincial CDC: Jian Su, Ran Tao, Ming Wu, Jie Yang, Jinyi Zhou, Yonglin Zhou. Suzhou CDC: Yihe Hu, Yujie Hua, Jianrong Jin, Fang Liu, Jingchao Liu, Yan Lu, Liangcai Ma, Aiyu Tang, Jun Zhang. **Qingdao** CDC: Liang Cheng, Ranran Du, Ruqin Gao, Feifei Li, Shanpeng Li, Yongmei Liu, Feng Ning, Zengchang Pang, Xiaohui Sun, Xiaocao Tian, Shaojie Wang, Yaoming Zhai, Hua Zhang, Licang CDC: Wei Hou, Silu Lv, Junzheng Wang. **Sichuan** Provincial CDC: Xiaofang Chen, Xianping Wu, Ningmei Zhang, Weiwei Zhou. Pengzhou CDC: Xiaofang Chen, Jianguo Li, Jiaqiu Liu, Guojin Luo, Qiang Sun, Xunfu Zhong. **Zhejiang** Provincial CDC: Weiwei Gong, Ruying Hu, Hao Wang,Meng Wang, Min Yu. Tongxiang CDC: Lingli Chen, Qijun Gu, Dongxia Pan，Chunmei Wang, Kaixu Xie, Xiaoyi Zhang.

### eFigure 1: Correlations of levels of 1,694 proteins between SomaScan-ANML and SomaScan-non-ANML

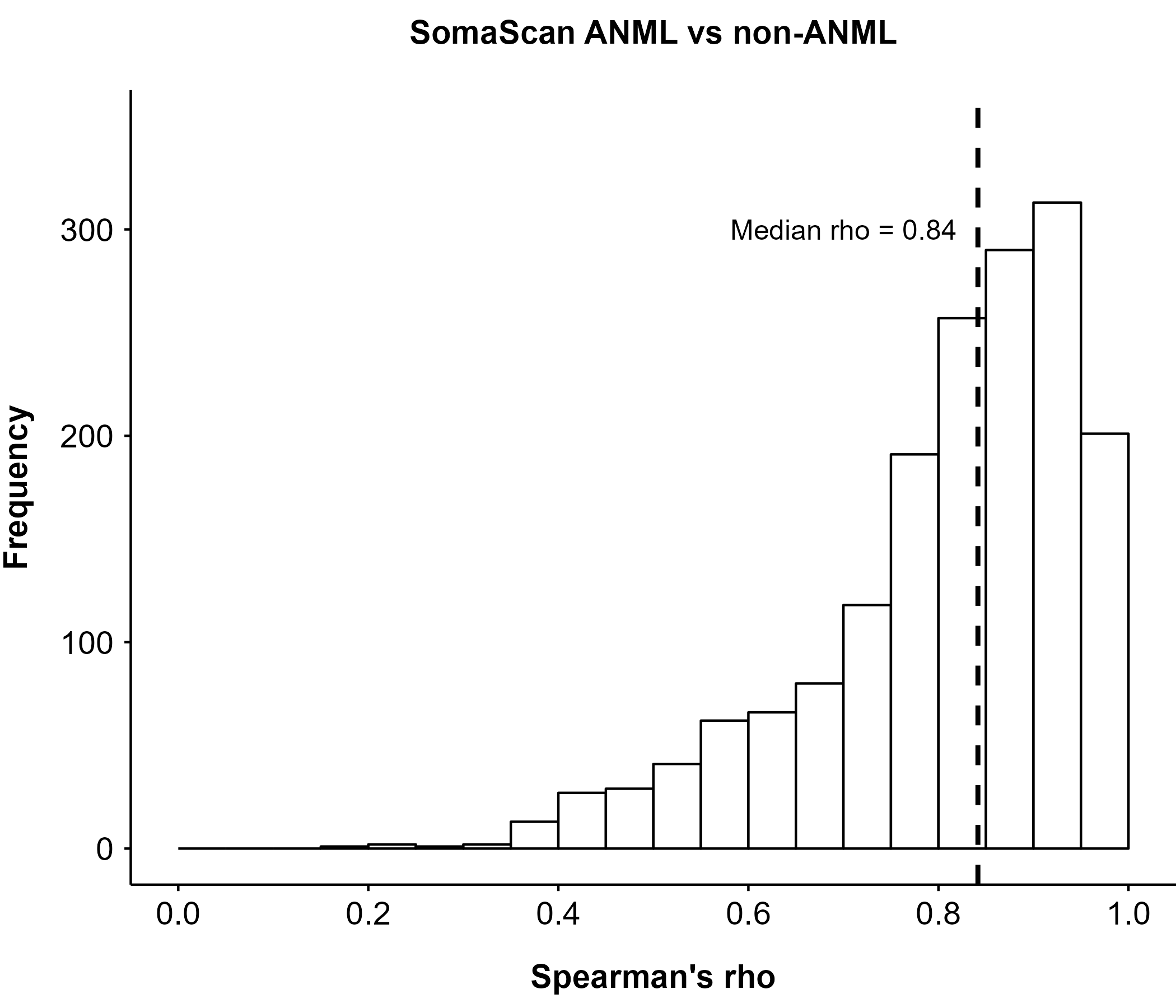

### eFigure 2: Correlations of levels of 1,694 proteins between OLINK and SomaScan in 2,025 subcohort participants

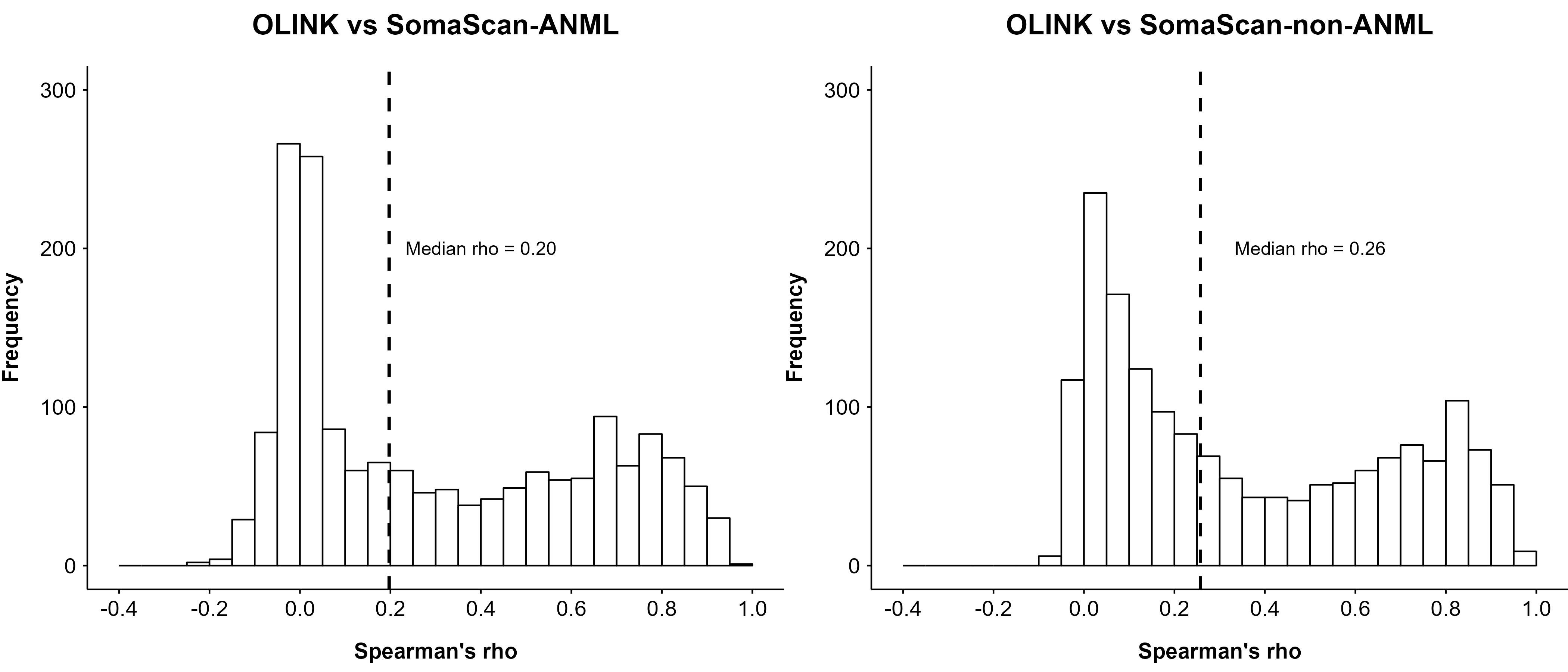

### eFigure 3: Pearson’s r between OLINK and SomaScan before and after log-transformation of SomaScan

Log-transformation (base-e) of the SomaScan data increased the median Pearson’s r with OLINK from 0.05/0.08 to 0.15/0.20 (for ANML/non-ANML).

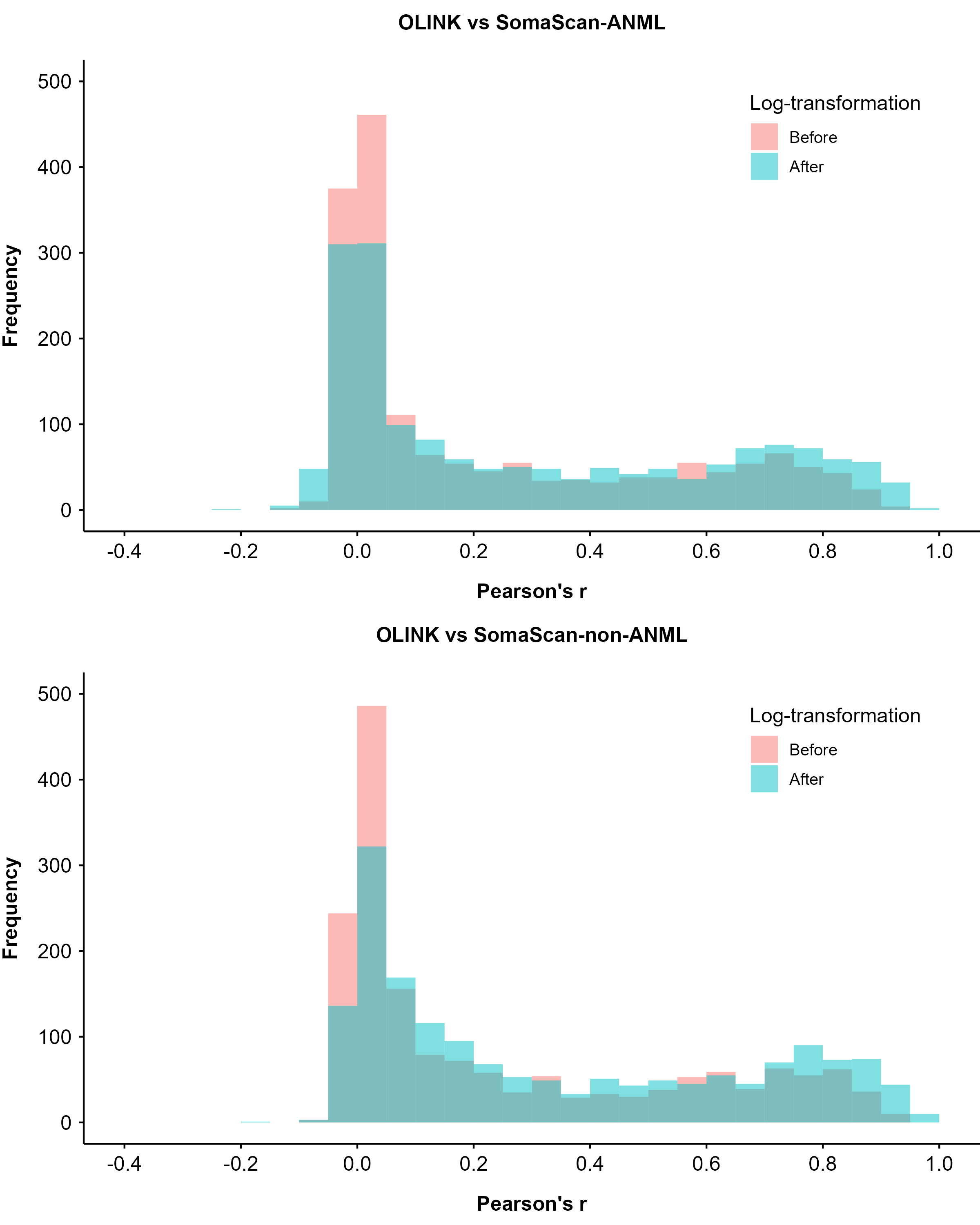

### eFigure 4: Correlations between protein levels measured by OLINK and SomaScan platforms according to degree of dilution in OLINK assay

Dashed line indicates the median rho.

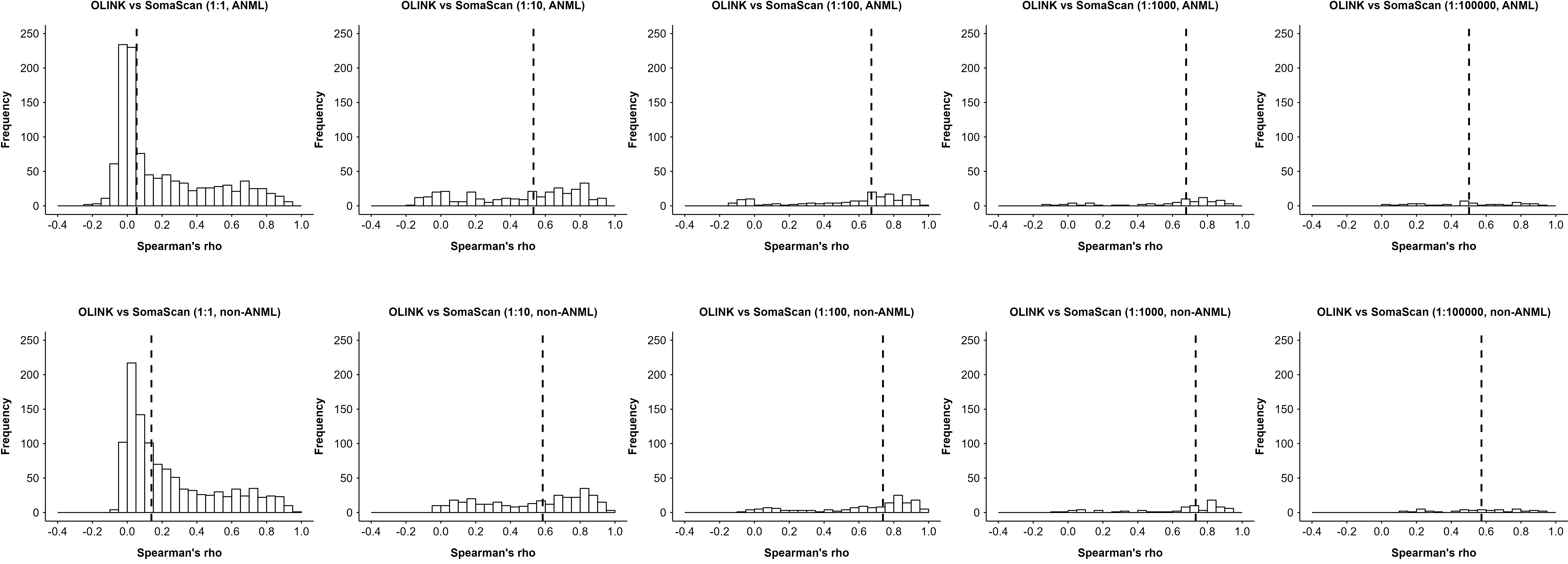

### eFigure 5: Correlations between protein levels measured by OLINK and SomaScan platforms according to degree of dilution in SomaScan assay

Dashed line indicates the median rho.

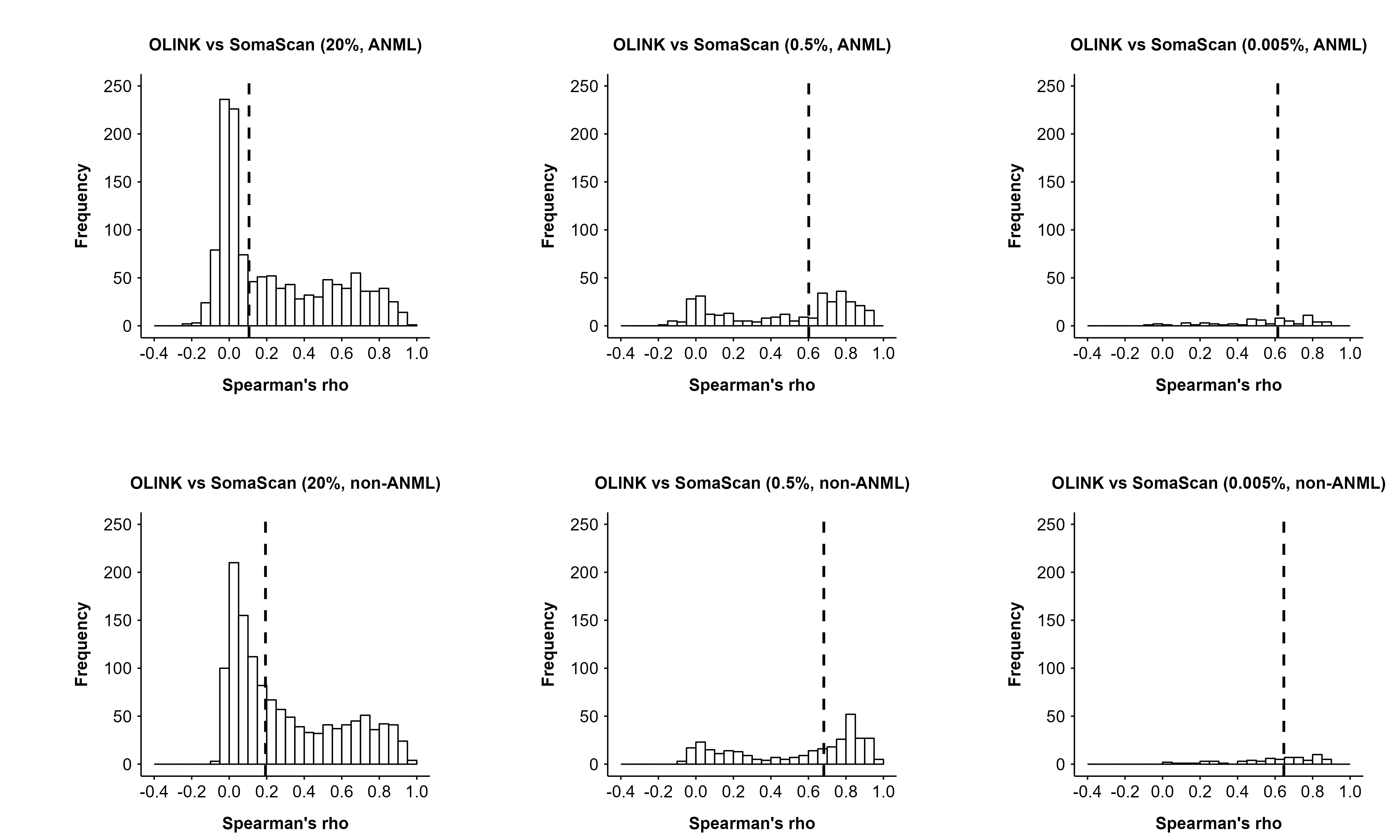

### eFigure 6: Comparison of –log10(p) of pQTLs between OLINK and SomaScan platforms

Comparison involves all sentinel pQTLs identified in one platform and the corresponding variant in the other platform.

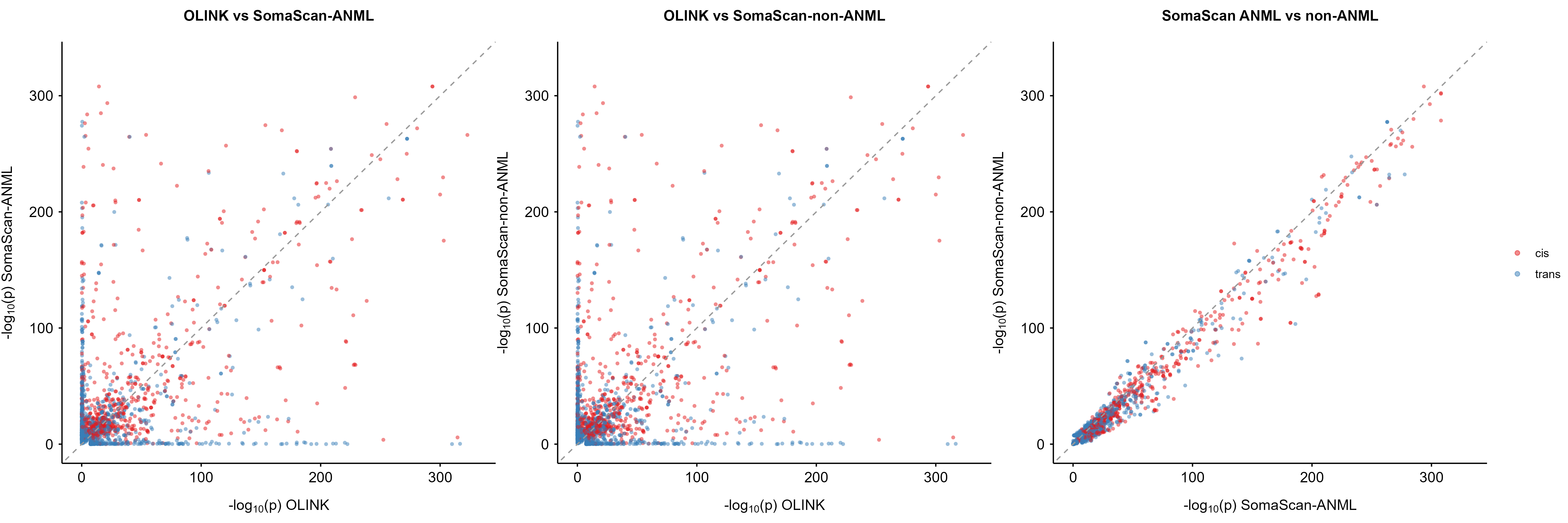

### eFigure 7: Number of proteins with *cis*-pQTLs discovered in SomaScan-ANML and SomaScan-non-ANML and colocalisation results

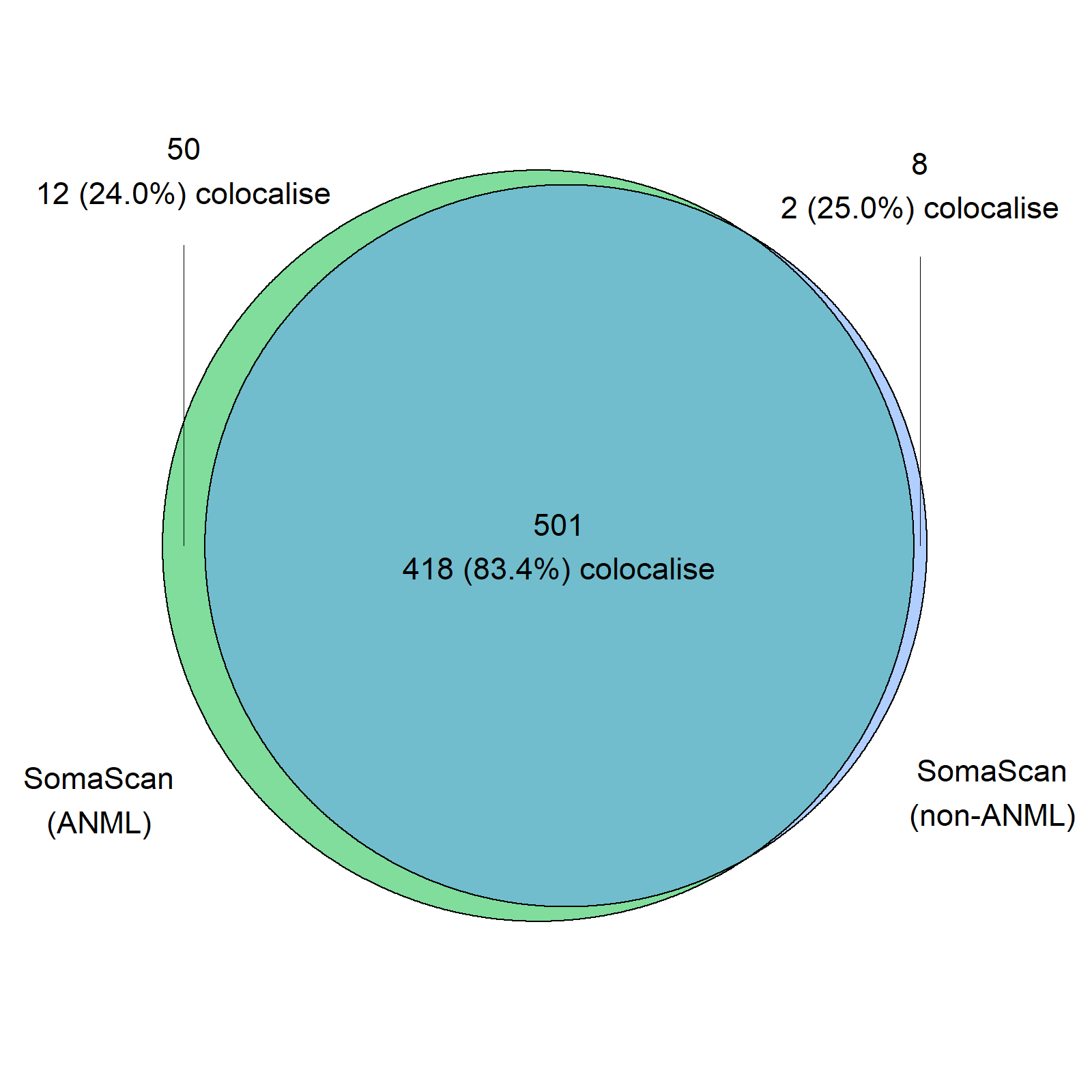

### eFigure 8: Number of proteins significantly associated with BMI and their effect sizes after applying Bonferroni correction for multiple testing

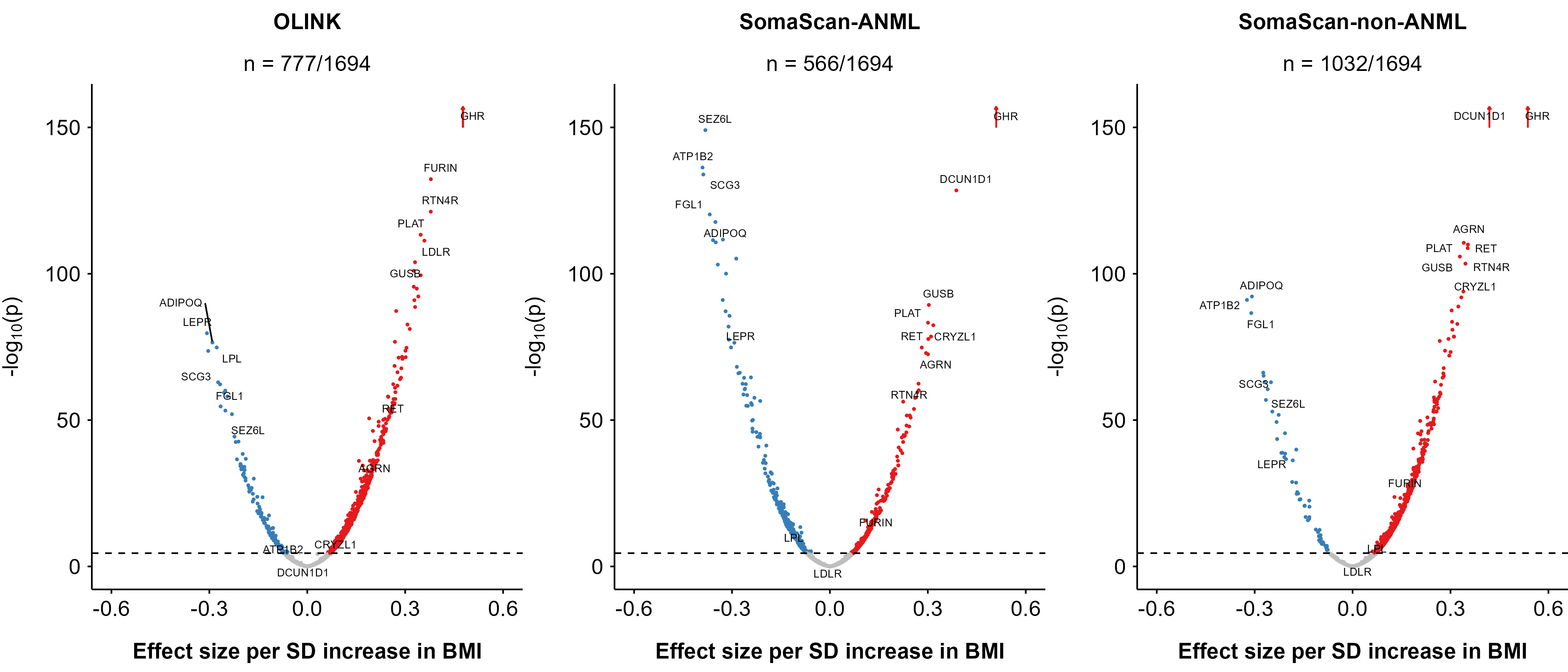

### eFigure 9: Comparison of effect sizes for proteins associated with BMI and risk of incident IHD between SomaScan ANML and non-ANML

Dark dots indicates shared associations between SomaScan-ANML and SomaScan-non-ANML, which were defined as significant associations found in both datasets that were also directionally consistent. Results were corrected using false discovery rate.

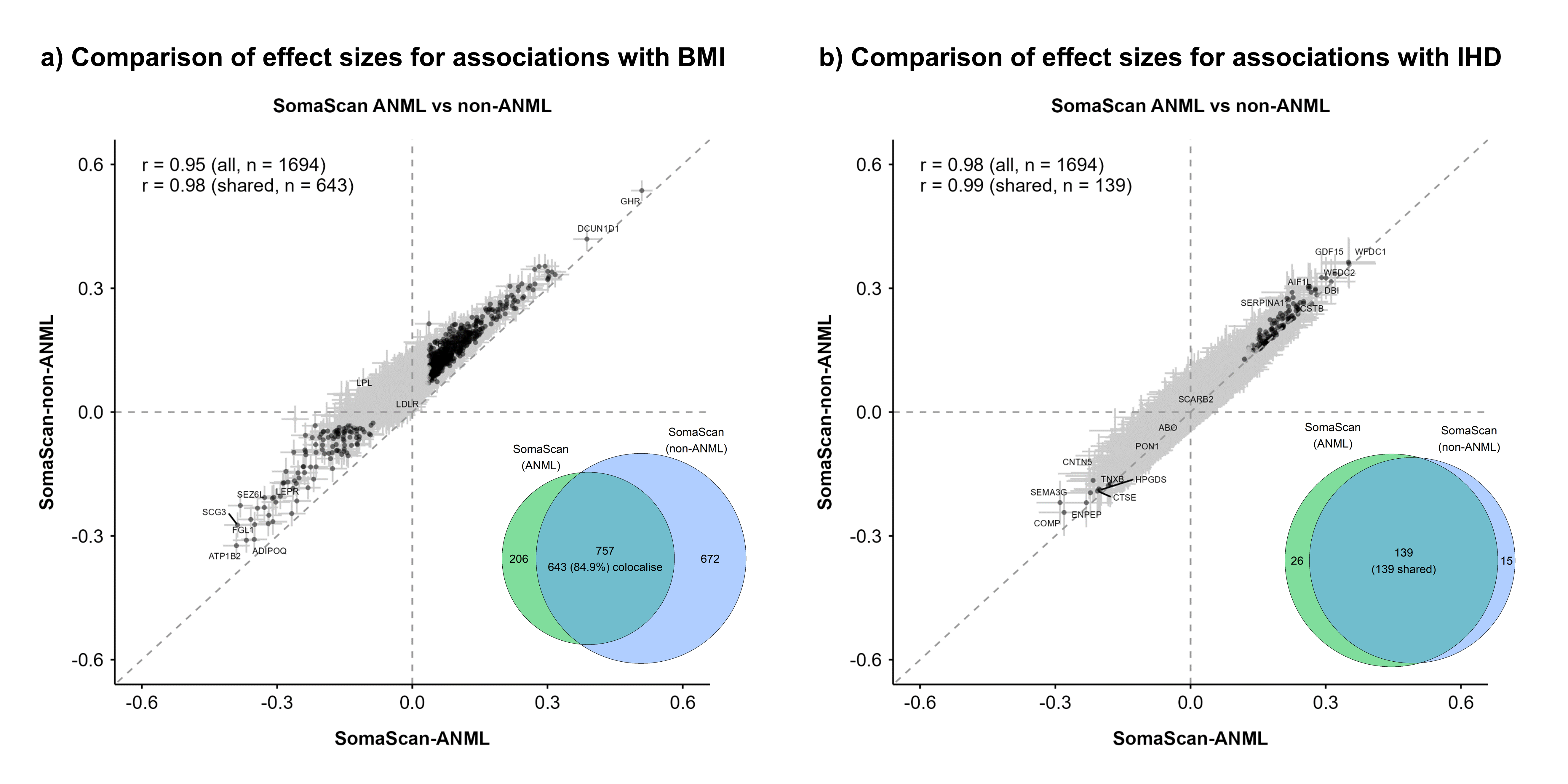

### eFigure 10: Number of proteins significantly associated with selected baseline characteristics for OLINK and SomaScan platforms

Results were corrected using false discovery rate within each trait and each platform. Analyses on ever regular smoking and regular vs occasional drinking were conducted in male participants only.

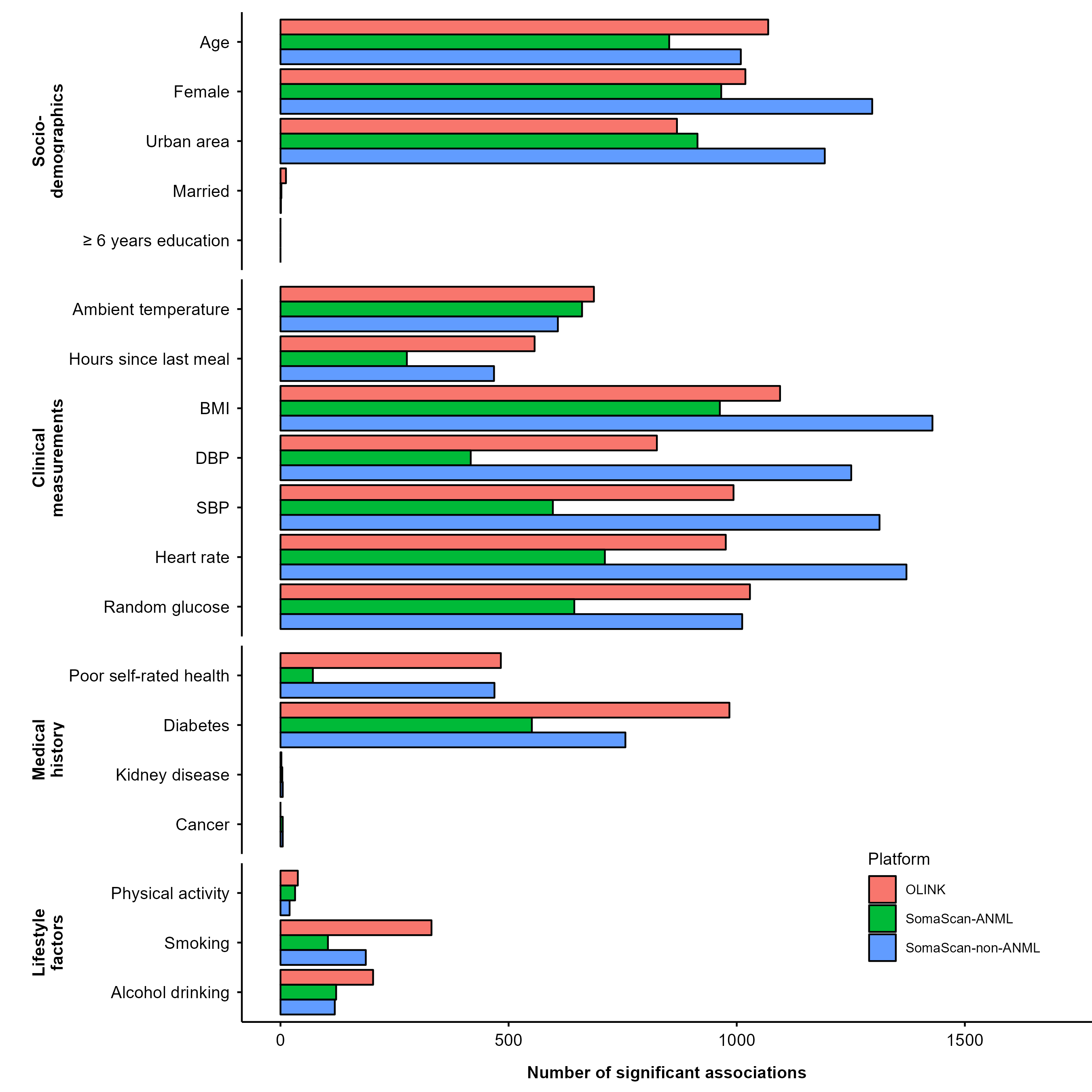

### eFigure 11: Concordance of associations of proteins with participant characteristics between OLINK and SomaScan platforms

Results were corrected using false discovery rate within each trait and each platform. Shared associations were defined as significant associations found in both platforms that were also directionally consistent. Results were corrected using false discovery rate. Analyses on ever regular smoking and regular vs occasional drinking were conducted in male participants only.

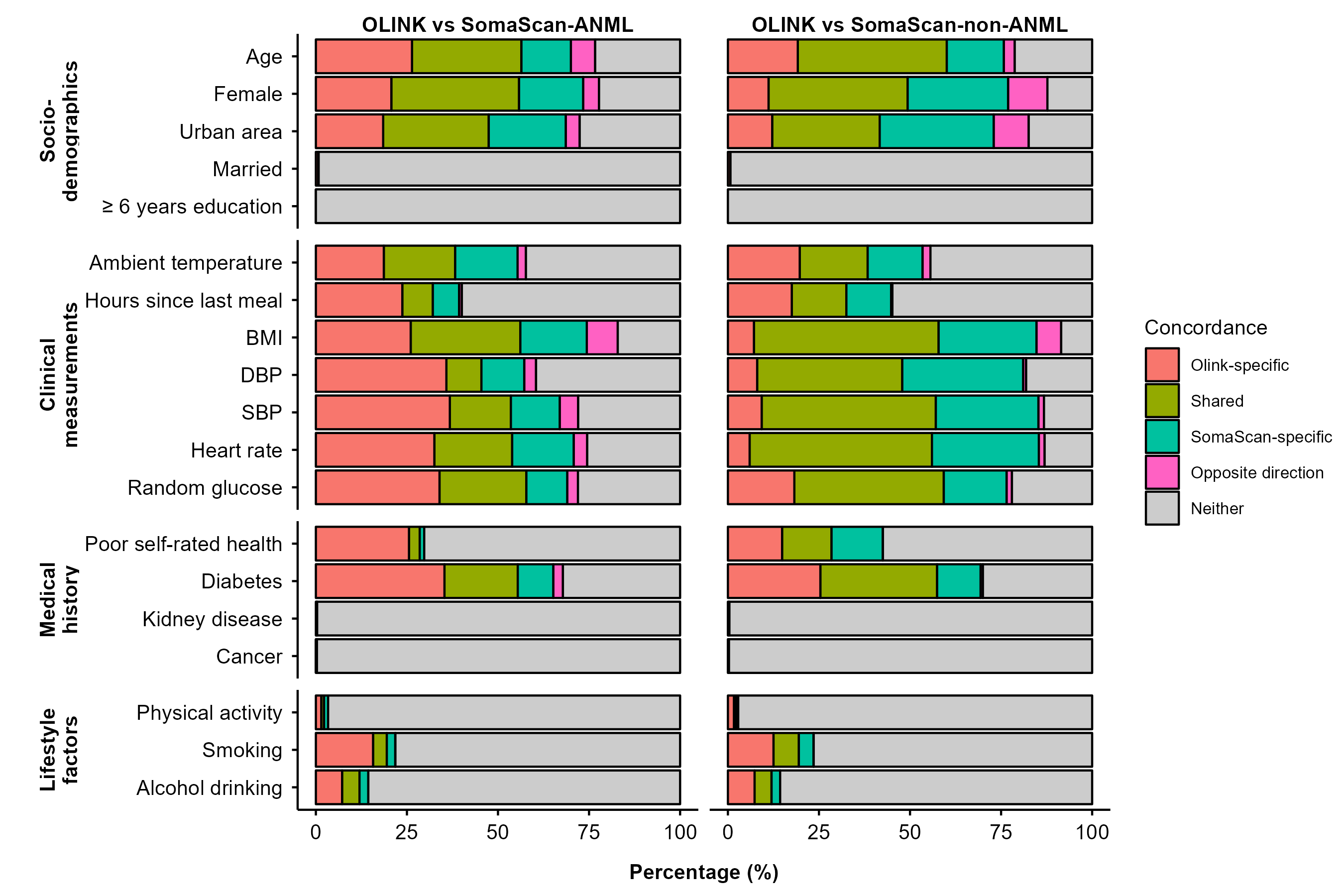

### eFigure 12: Concordance of associations of proteins with participant characteristics between SomaScan ANML and non-ANML

Results were corrected using false discovery rate within each trait and each platform. Shared associations were defined as significant associations found in both datasets that were also directionally consistent. Analyses on ever regular smoking and regular vs occasional drinking were conducted in male participants only.

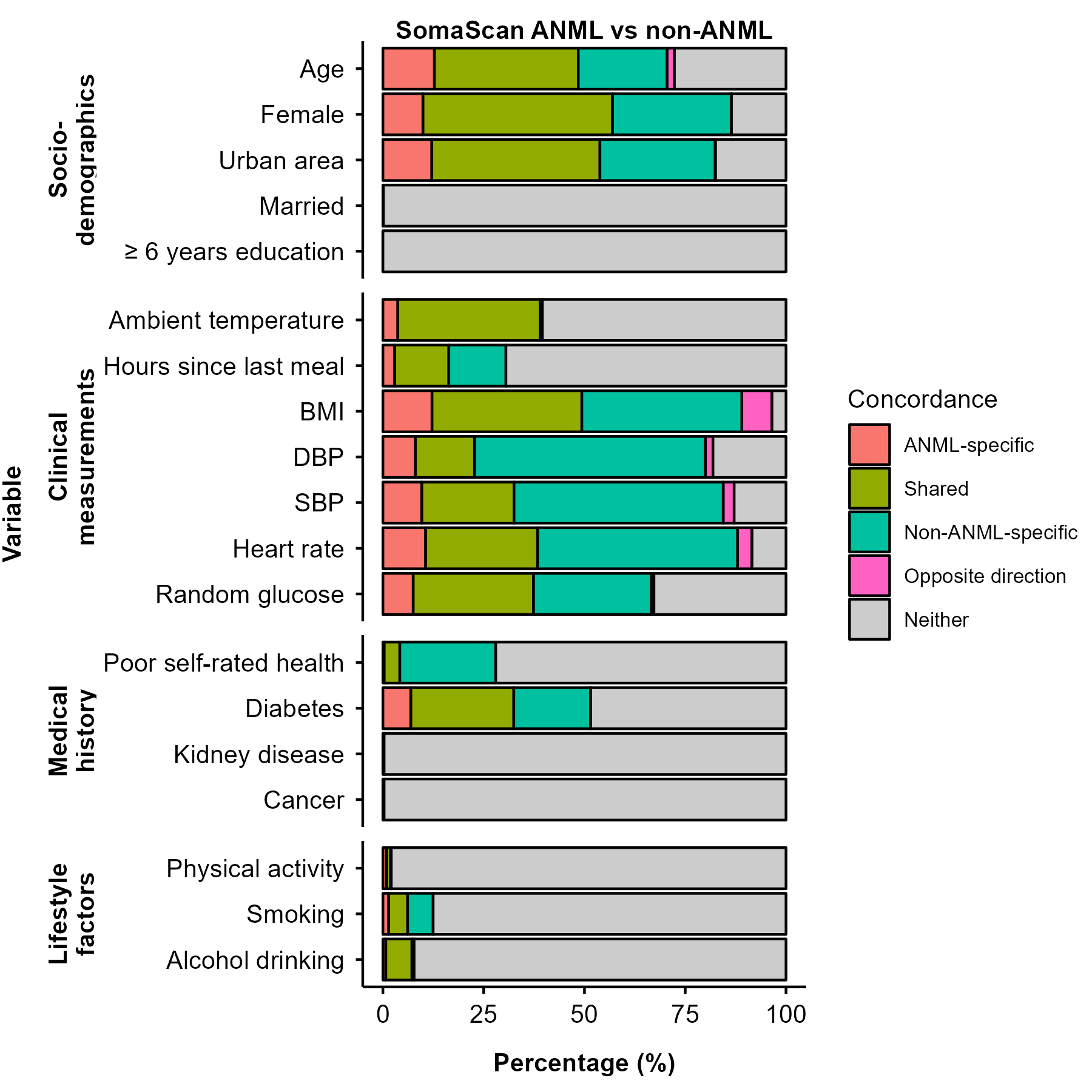

### eFigure 13: Correlation of effect sizes for proteins significantly associated with participant characteristics between OLINK and SomaScan platforms

Results were corrected using false discovery rate within each trait and each platform. Shared associations were defined as significant associations found in both platforms that were also directionally consistent. Analyses on ever regular smoking and regular vs occasional drinking were conducted in male participants only.

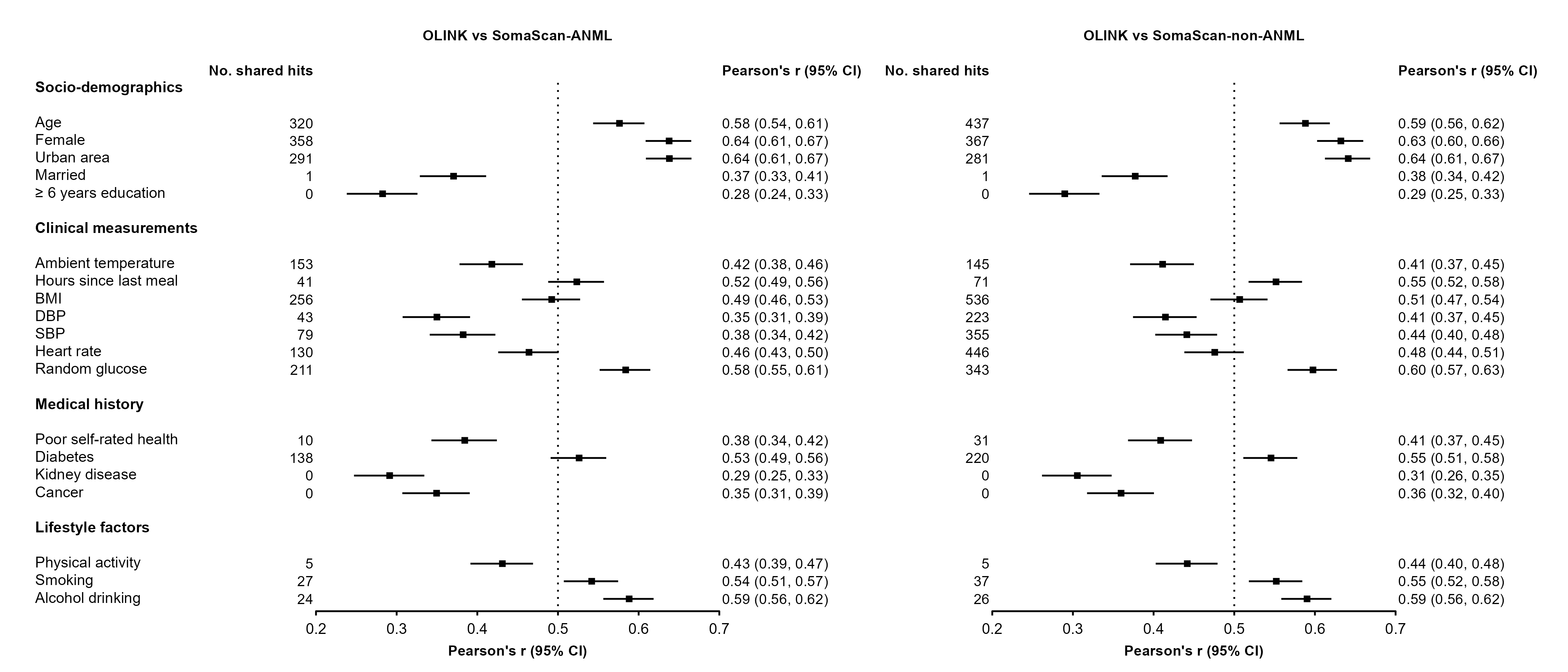

### eFigure 14: Correlation of effect sizes for proteins significantly associated with participant characteristics between SomaScan ANML and non-ANML

Results were corrected using false discovery rate within each trait and each platform. Shared associations were defined as significant associations found in both datasets that were also directionally consistent. Analyses on ever regular smoking and regular vs occasional drinking were conducted in male participants only.

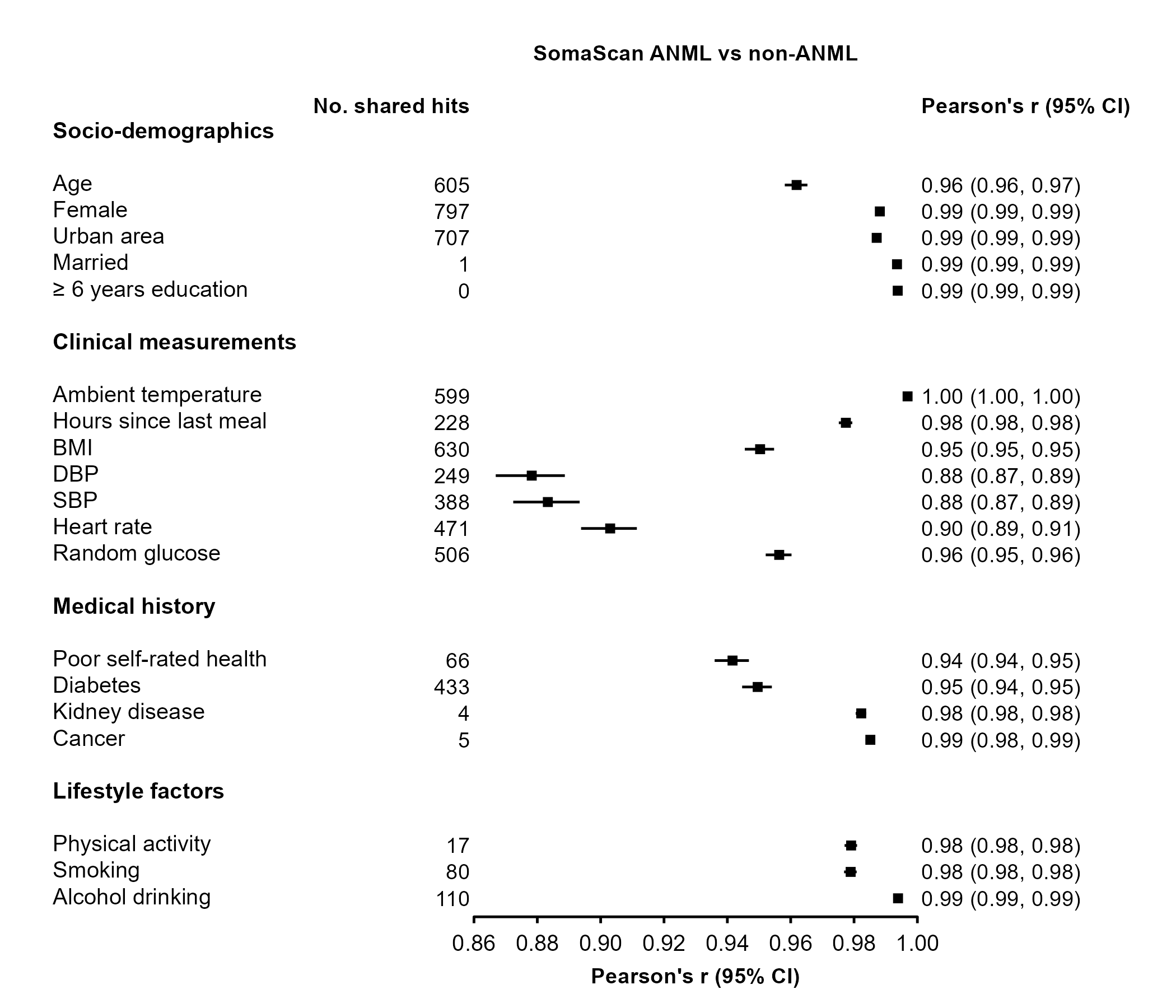

### eFigure 15: Number of proteins significantly associated with participant characteristics for OLINK and SomaScan platforms, in subcohort participants only

Results were corrected using false discovery rate within each trait and each platform. Analyses on ever regular smoking and regular vs occasional drinking were conducted in male participants only.

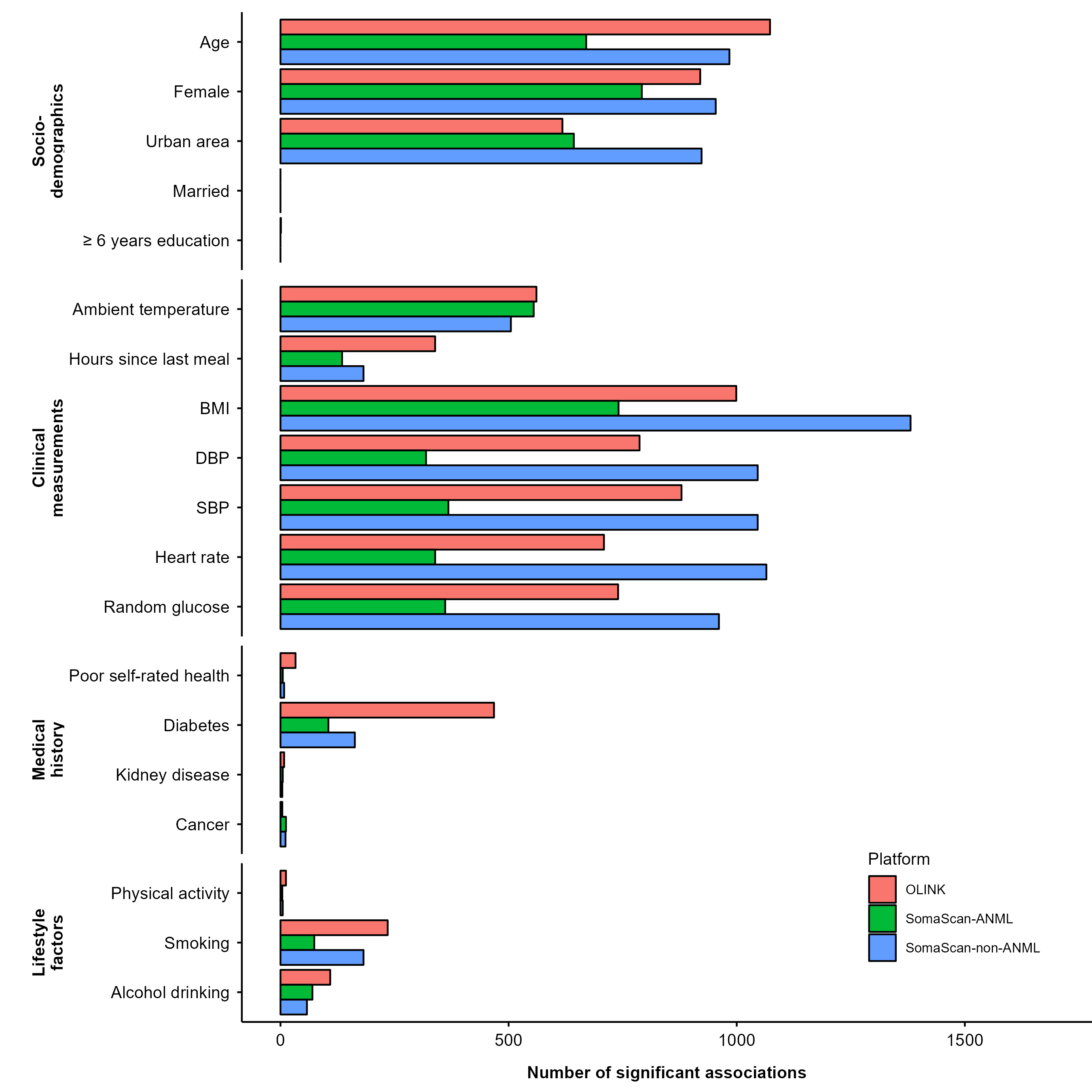

### eFigure 16: Concordance of associations of proteins with participant characteristics between OLINK and SomaScan platforms, in subcohort participants only

Results were corrected using false discovery rate within each trait and each platform. Shared associations were defined as significant associations found in both platforms that were also directionally consistent. Analyses on ever regular smoking and regular vs occasional drinking were conducted in male participants only.

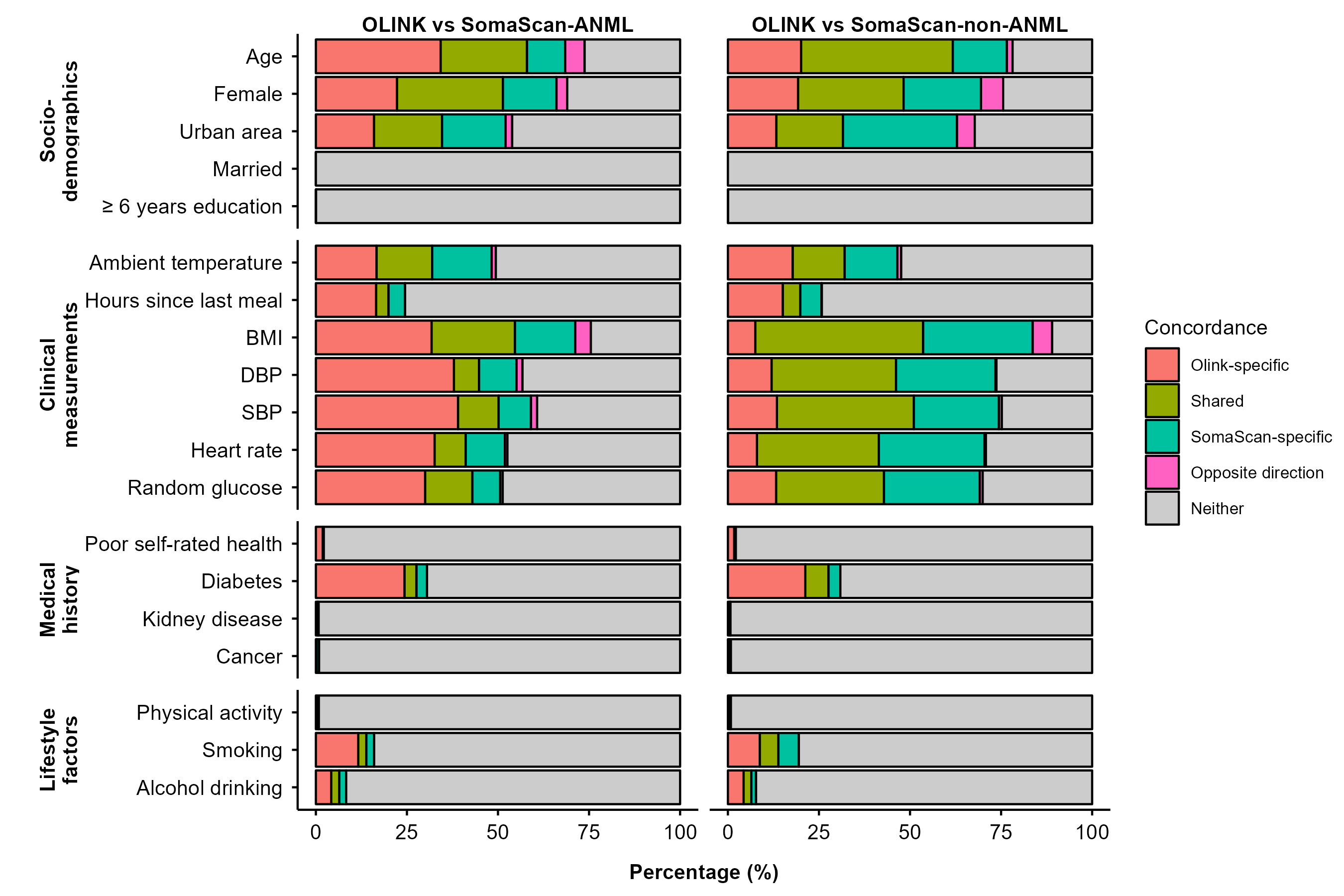

### eFigure 17: Concordance of associations of proteins with participant characteristics between SomaScan ANML and non-ANML, in subcohort participants only

Results were corrected using false discovery rate within each trait and each platform. Shared associations were defined as significant associations found in both datasets that were also directionally consistent. Analyses on ever regular smoking and regular vs occasional drinking were conducted in male participants only.

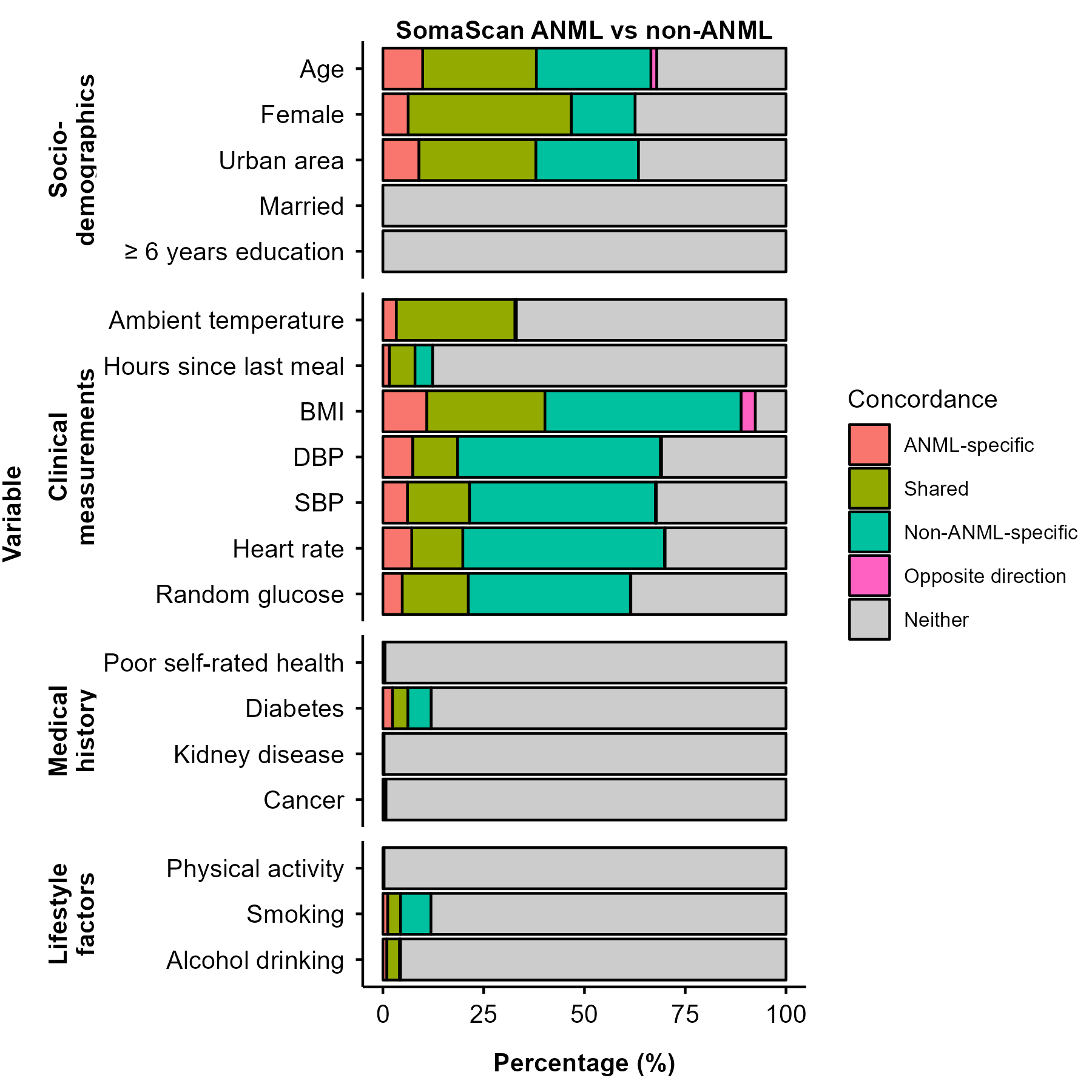

### eFigure 18: Correlation of effect sizes for proteins significantly associated with participant characteristics between OLINK and SomaScan platforms, in subcohort participants only

Results were corrected using false discovery rate within each trait and each platform. Shared associations were defined as significant associations found in both platforms that were also directionally consistent. Analyses on ever regular smoking and regular vs occasional drinking were conducted in male participants only.

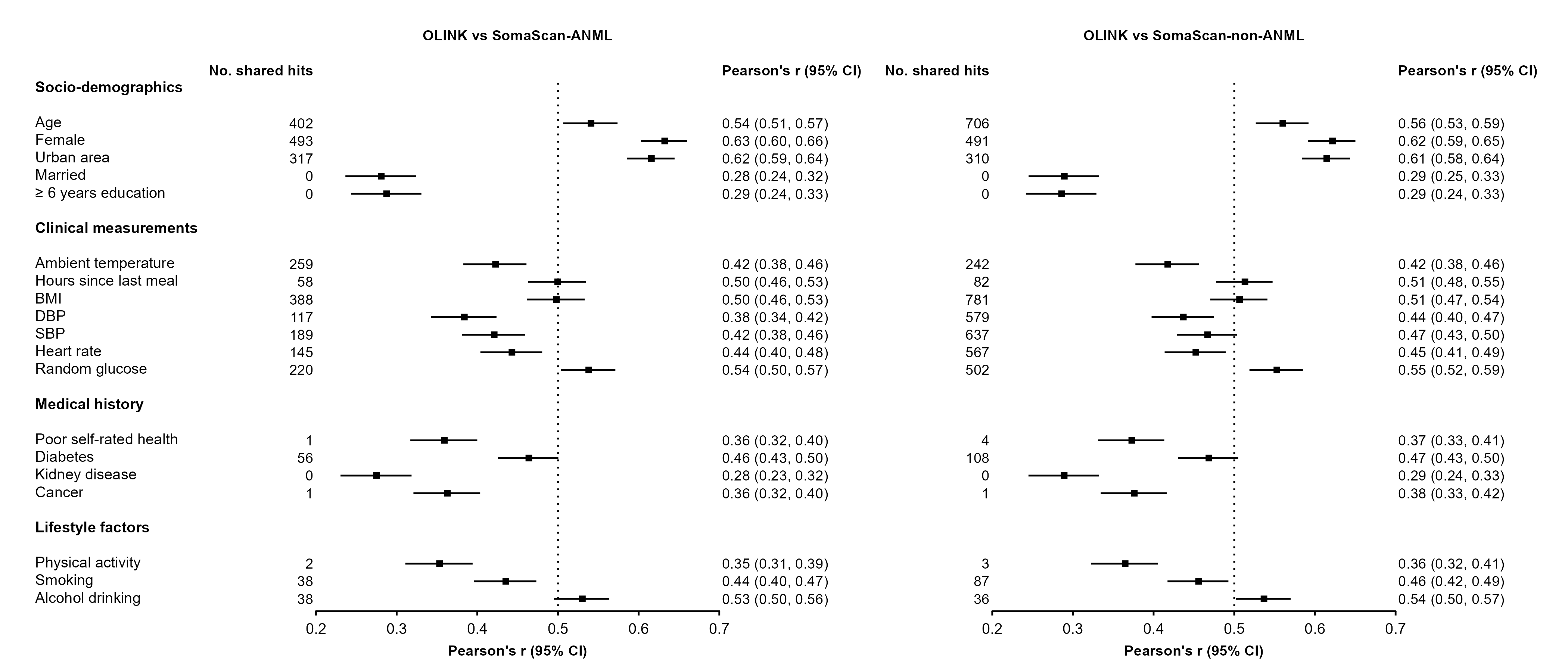

### eFigure 19: Correlation of effect sizes for proteins significantly associated with participant characteristics between SomaScan ANML and non-ANML, in subcohort participants only

Results were corrected using false discovery rate within each trait and each platform. Shared associations were defined as significant associations found in both datasets that were also directionally consistent. Analyses on ever regular smoking and regular vs occasional drinking were conducted in male participants only.

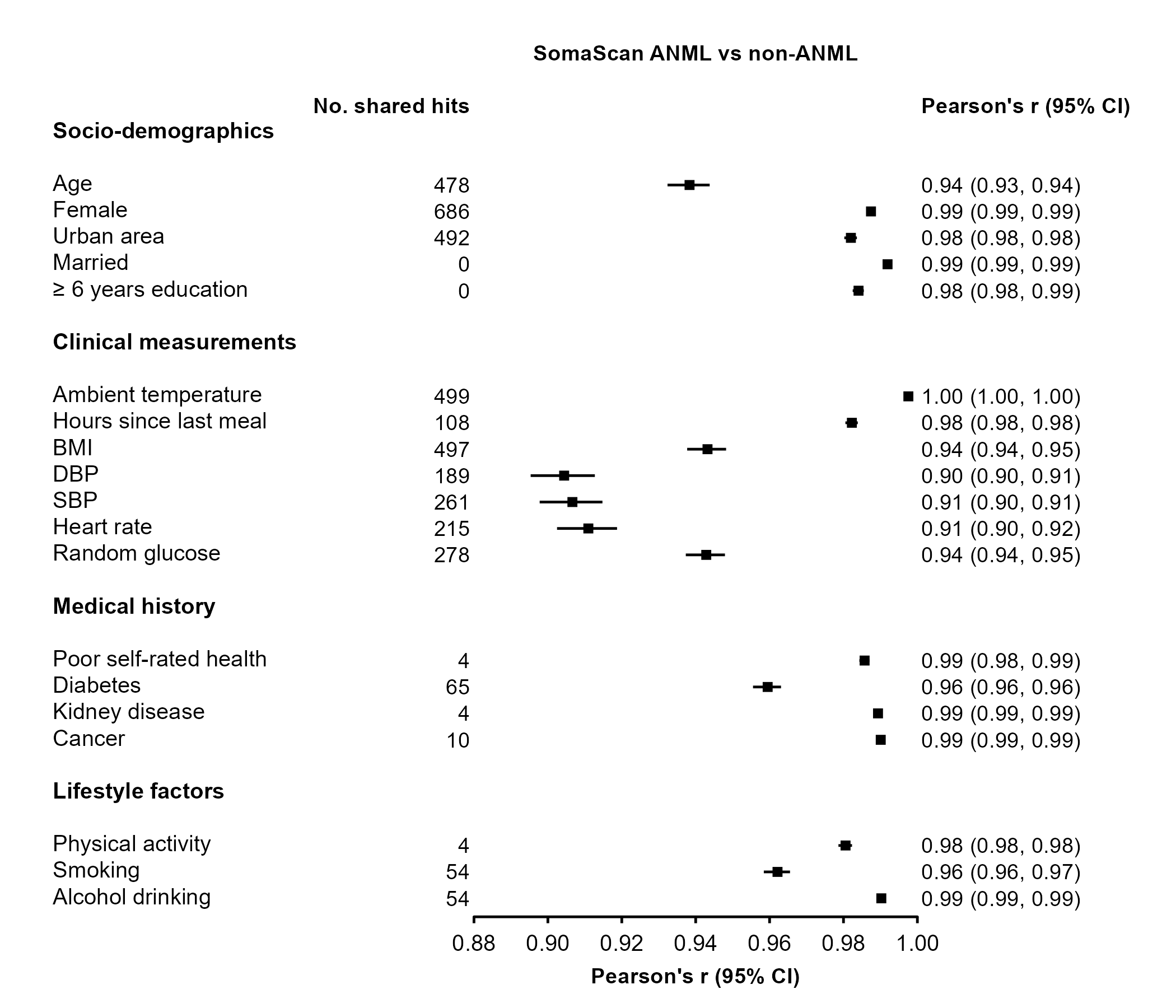

### eFigure 20: Number of proteins significantly associated with incident IHD and their effect sizes after applying Bonferroni correction for multiple testing

Raw p-values shown, with dashed lines indicating Bonferroni corrected significant thresholds.

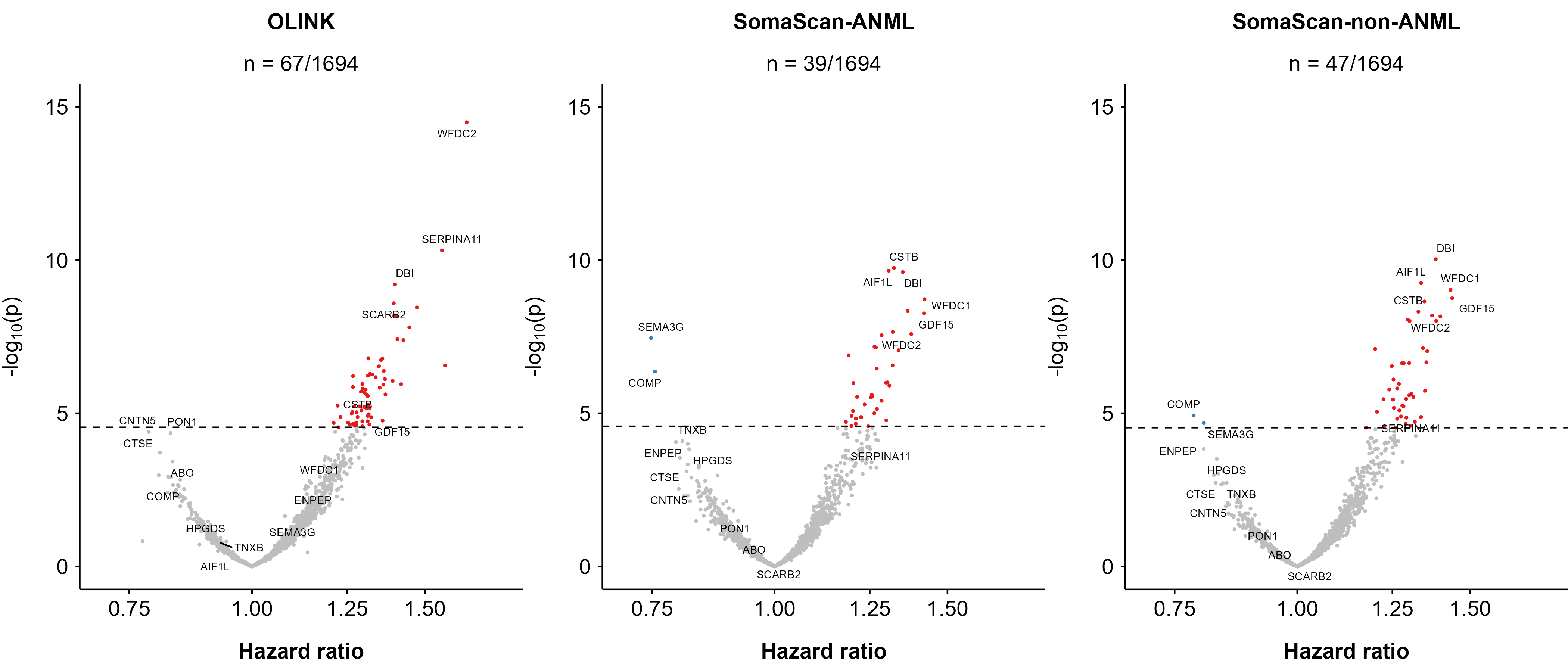

### eFigure 21: Correlations between protein levels measured by SOMAmers targeting the same protein

Analyses conducted on 472 proteins targeted by 2 to 9 SOMAmers (1,037 unique SOMAmers in total).

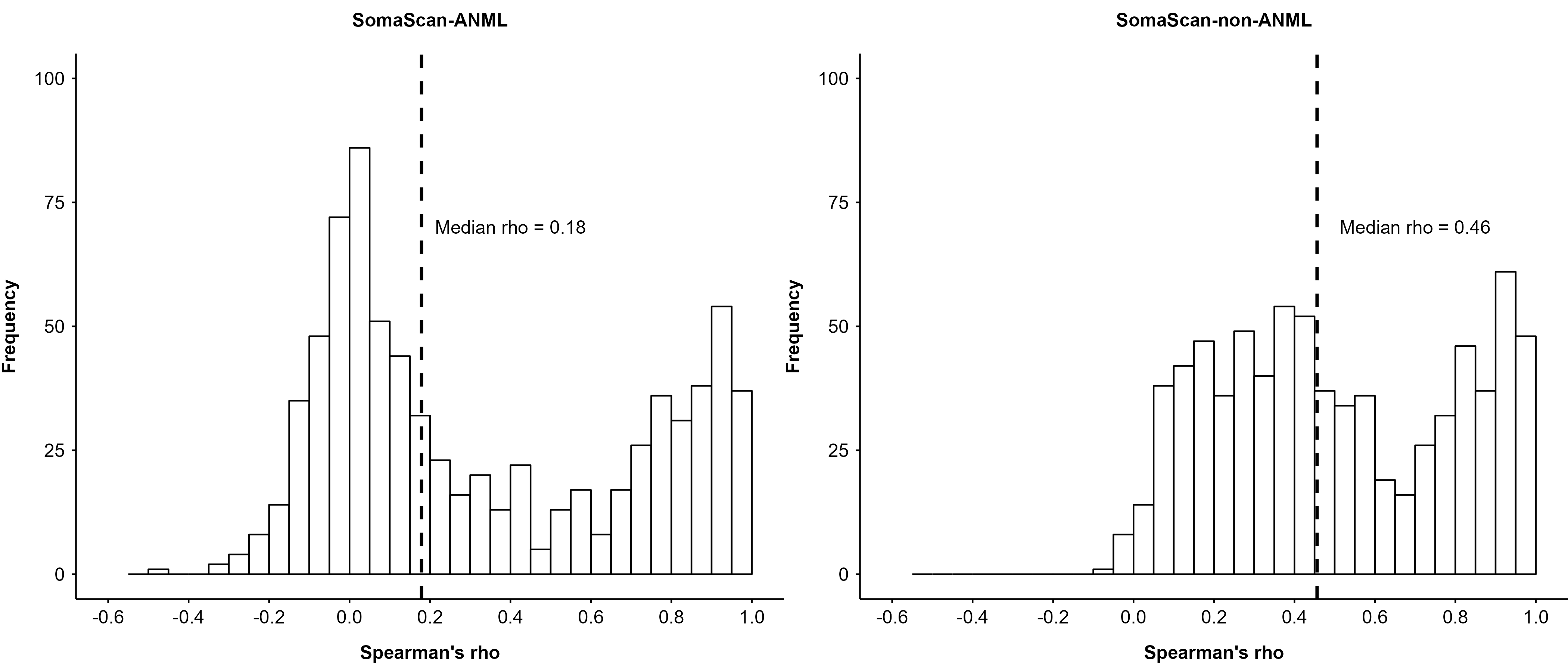

### eFigure 22: Correlations between protein levels measured by OLINK-SomaScan reagent pairs involving multiple SOMAmers

Analyses conducted on 472 proteins targeted by one OLINK reagent and 2 to 9 SOMAmers (1,037 unique SOMAmers in total), constituting 1,053 OLINK-SomaScan reagent pairs.

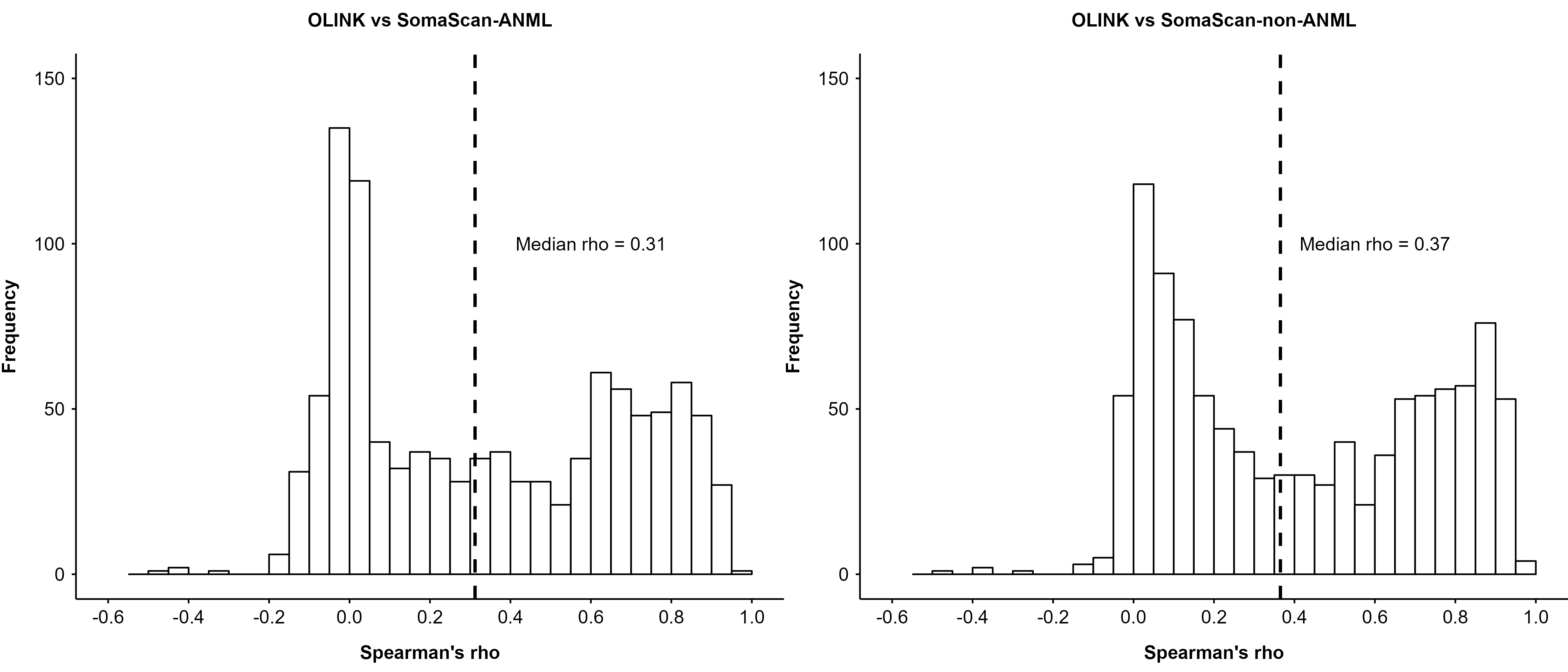

### eFigure 23: Correlations between protein levels measured by OLINK reagents targeting the same protein

Analyses conducted on 18 proteins targeted by 2 OLINK reagents (22 unique OLINK reagents in total).

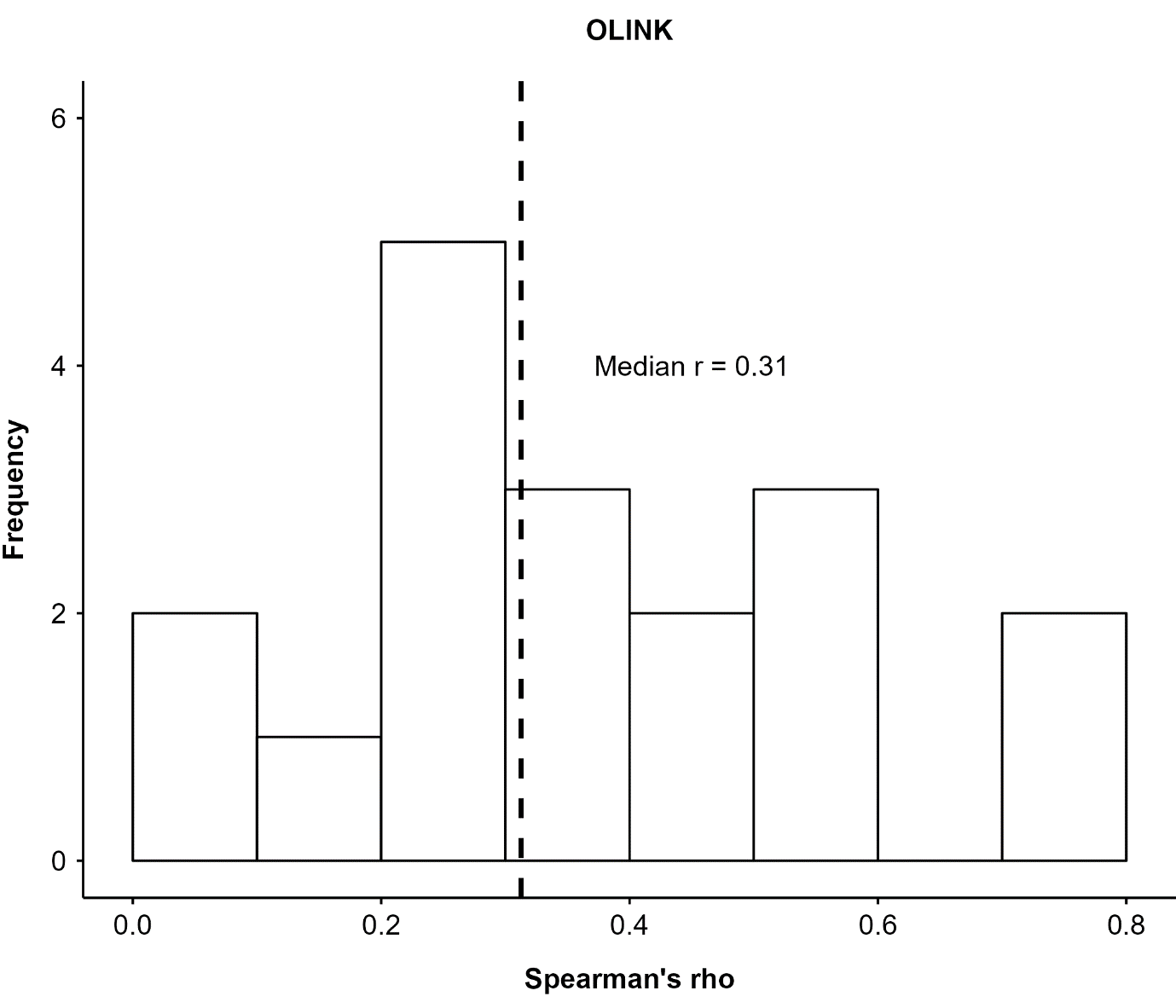

### eFigure 24: Correlations between protein levels measured by OLINK-SomaScan reagent pairs involving multiple OLINK reagents

Analysis conducted on 18 proteins targeted by one SOMAmer and also 2 OLINK reagents (22 unqiue OLINK reagents in total), constituting 36 OLINK-SomaScan reagent pairs.

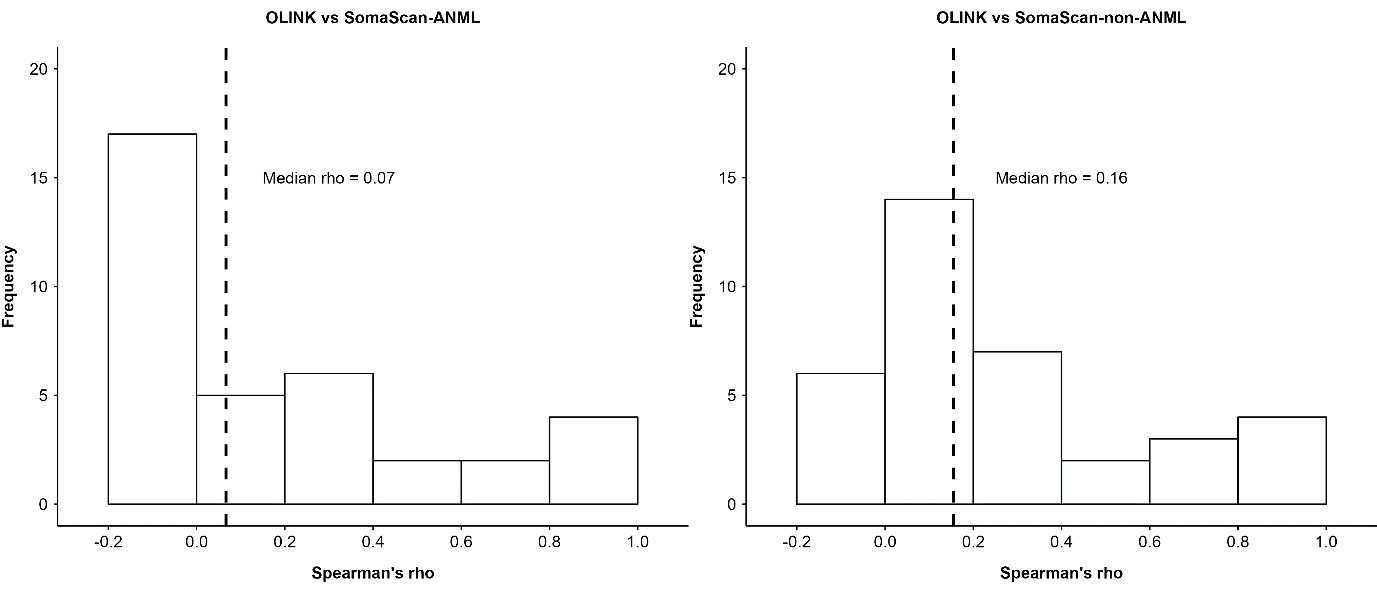

### eFigure 25: Colocalisation of *cis*-pQTLs for ALDH2 measured by OLINK and SomaScan platforms

CS: Credible sets from fine mapping, with 1 indicating results from OLINK and 2 from SomaScan (ANML). PIP: Posterior inclusion probability (PIP) from fine mapping. PP: Posterior probability from colocalisation.

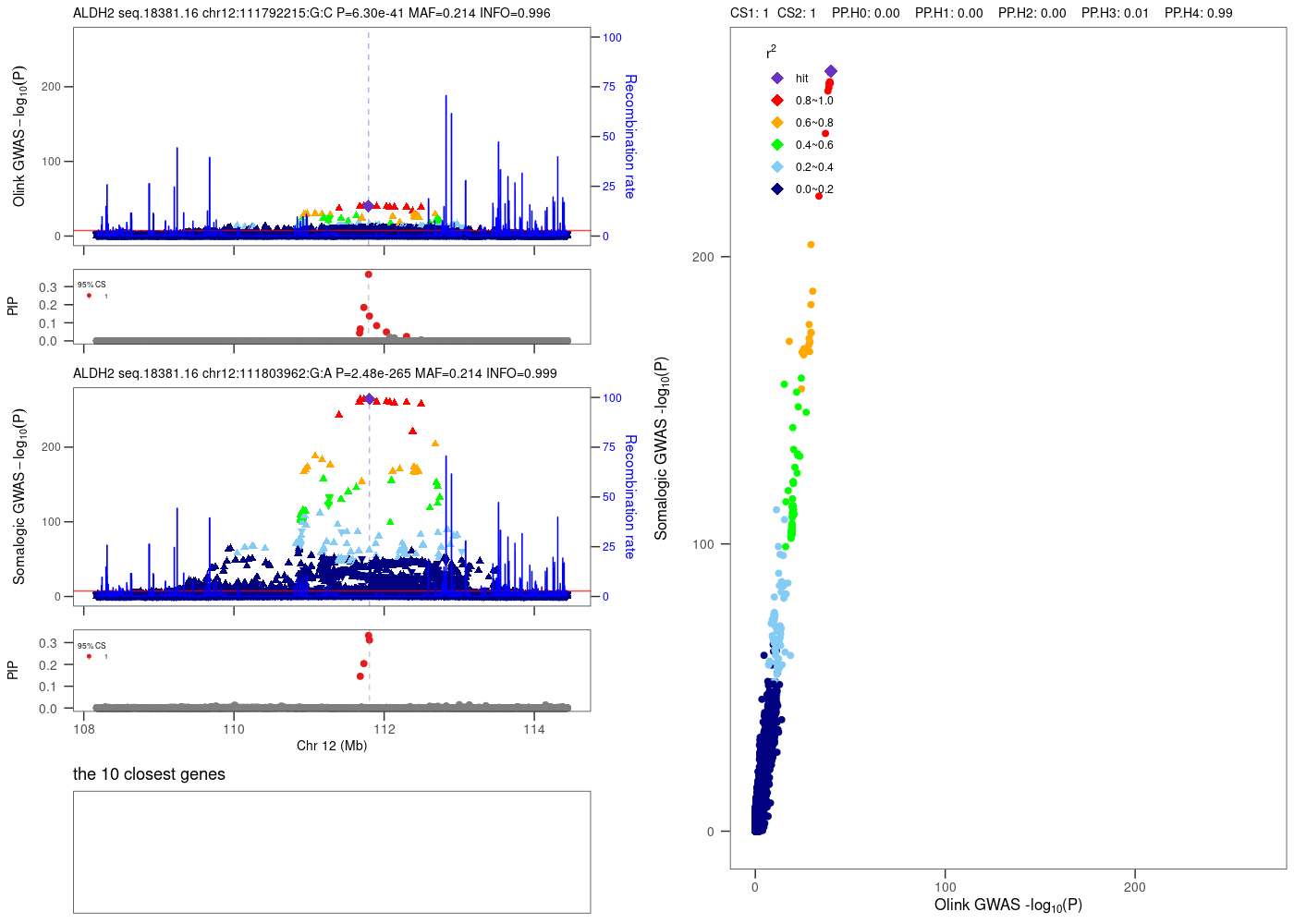

### eFigure 26: Colocalisation of *cis*-pQTLs for PLA2G7 measured by OLINK and SomaScan platforms

CS: Credible sets from fine mapping, with 1 indicating results from OLINK and 2 from SomaScan (ANML). PIP: Posterior inclusion probability (PIP) from fine mapping. PP: Posterior probability from colocalisation.

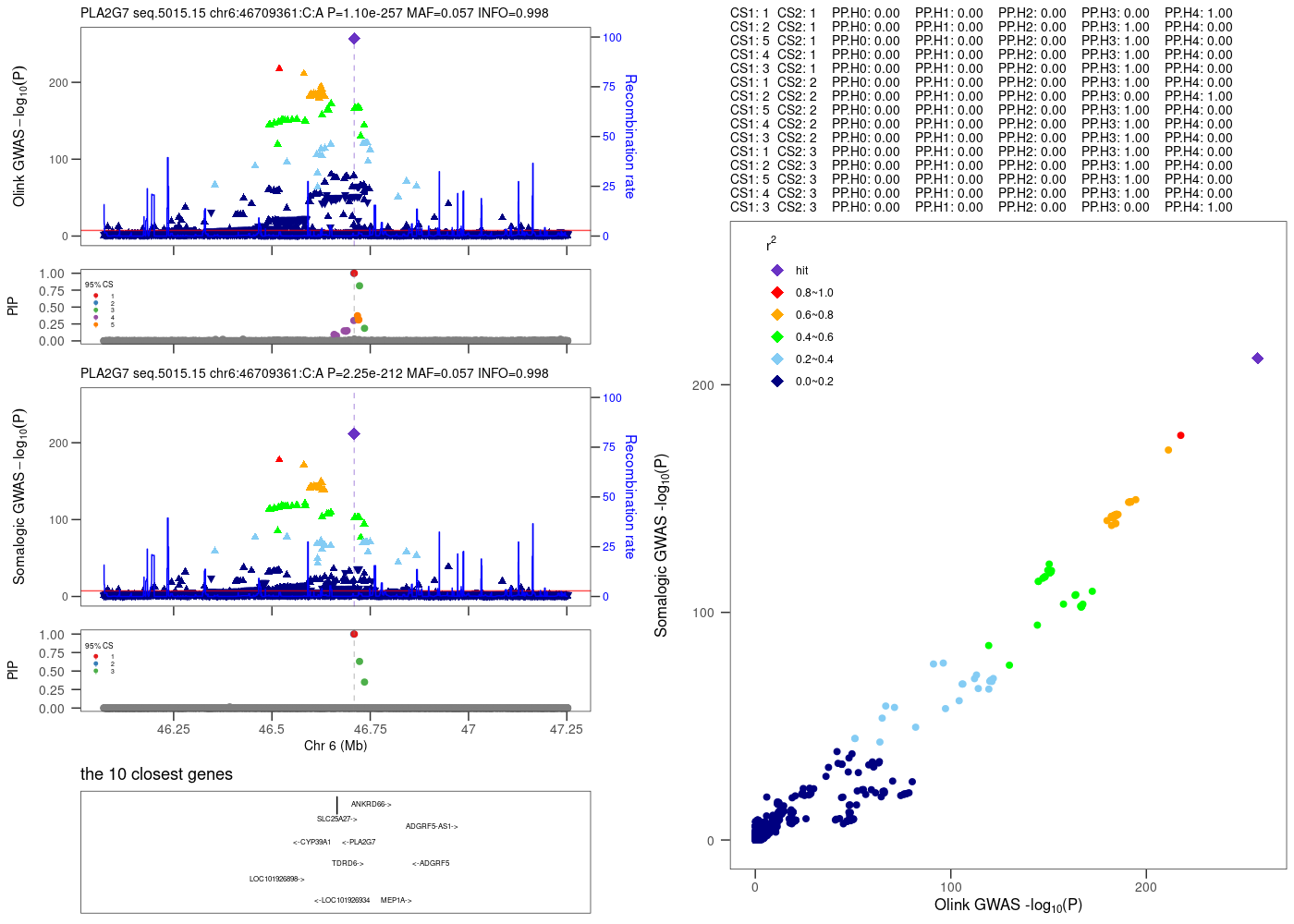

### eFigure 27: An example of colocalisation of *cis*-pQTLs between a SOMAmer targeting TNC and the corresponding OLINK reagent

CS: Credible sets from fine mapping, with 1 indicating results from OLINK and 2 from SomaScan (ANML). PIP: Posterior inclusion probability (PIP) from fine mapping. PP: Posterior probability from colocalisation.

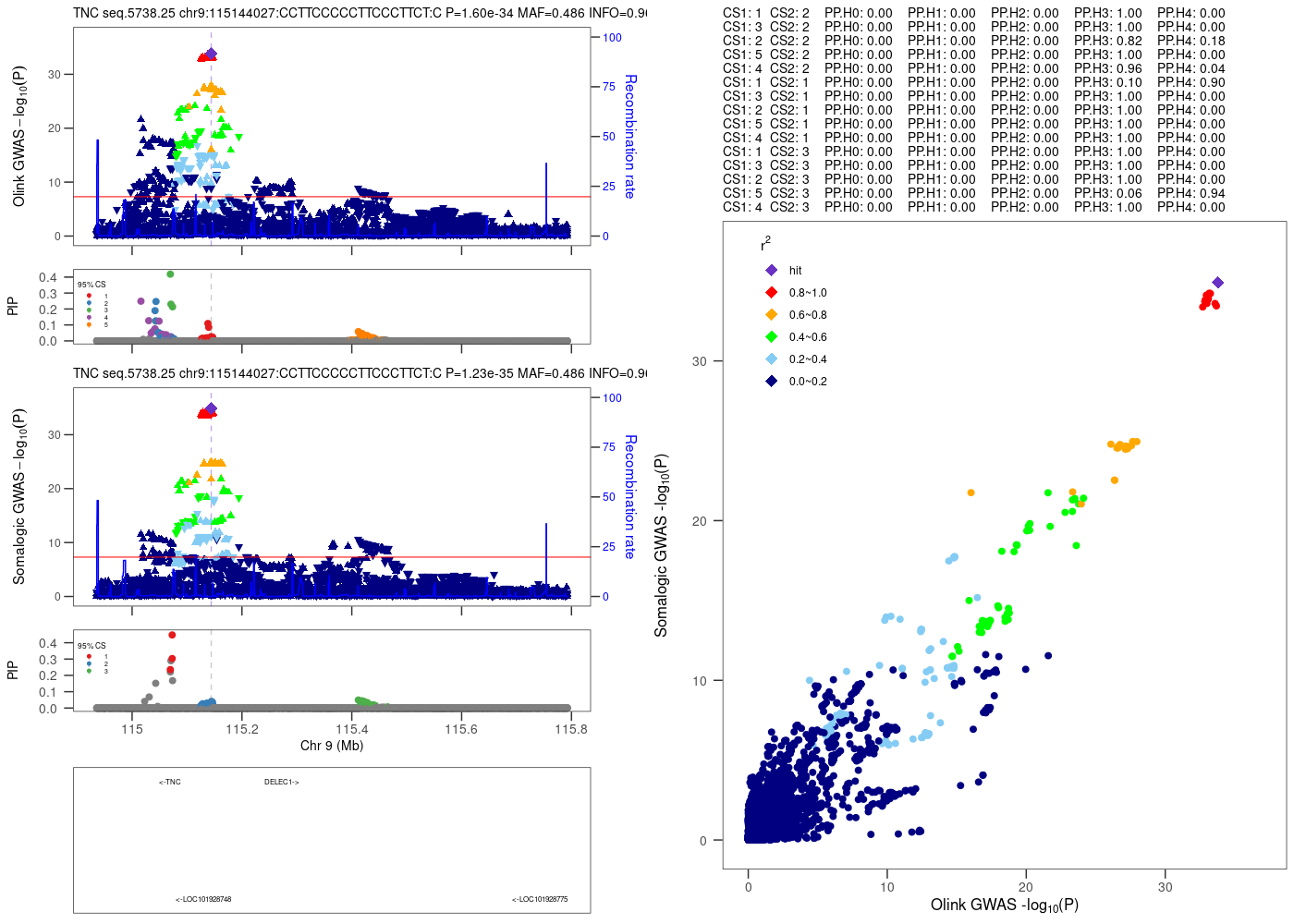

### eFigure 28: An example of no colocalisation of *cis*-pQTLs between a SOMAmer targeting TNC and the corresponding OLINK reagent

CS: Credible sets from fine mapping, with 1 indicating results from OLINK and 2 from SomaScan (ANML). PIP: Posterior inclusion probability (PIP) from fine mapping. PP: Posterior probability from colocalisation.

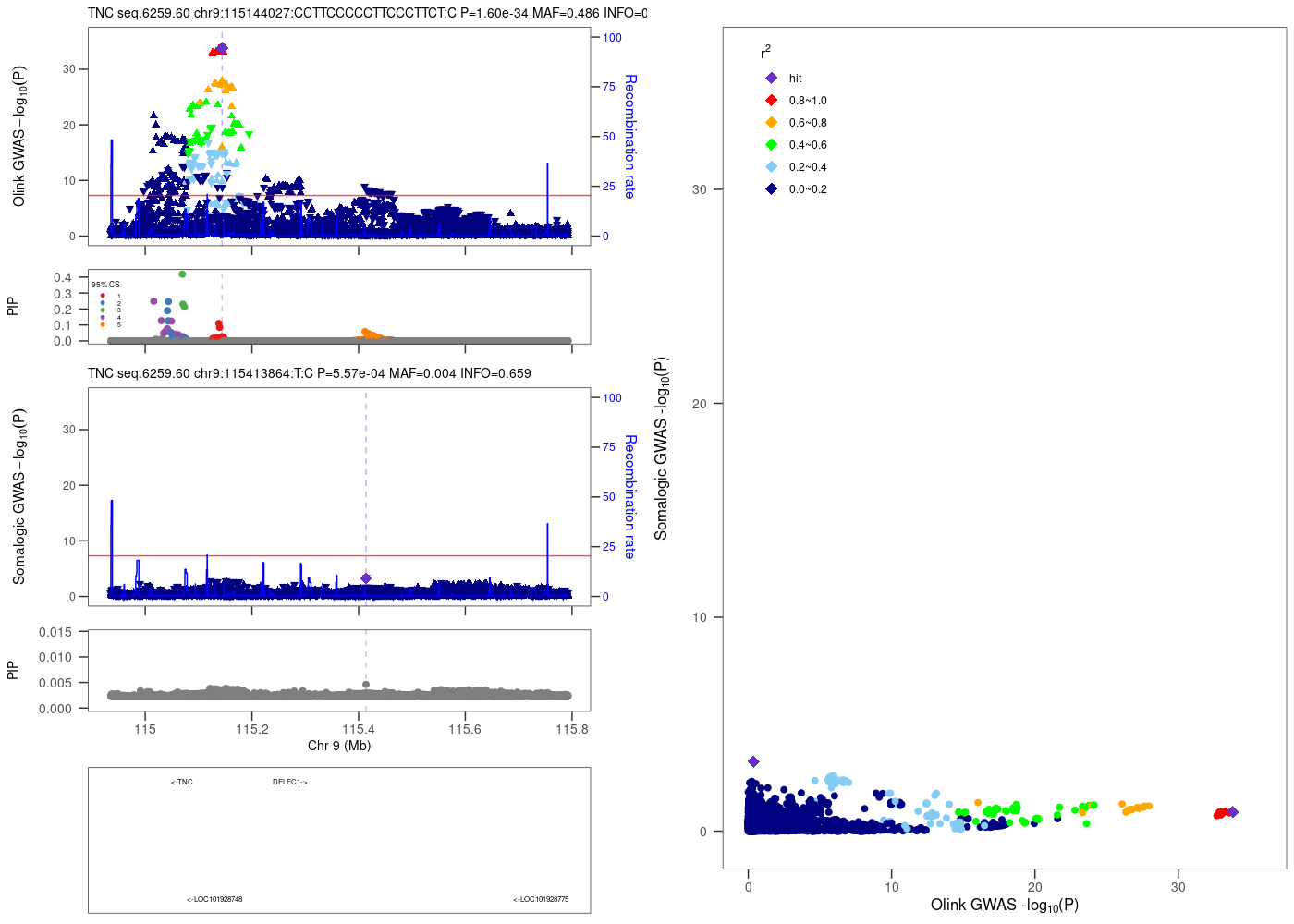

### eTable 1: Protein annotations retrieved from UniProt Knowledgebase for 1,694 overlapping proteins

| **Protein characteristic** | **Frequency** | **Percentage** |
| --- | --- | --- |
| Initiator methionine | 173 | 10.2 |
| Signal peptide | 1,007 | 59.4 |
| Transit peptide | 41 | 2.4 |
| Propeptide | 252 | 14.9 |
| Peptide | 55 | 3.2 |
| Topological domain | 504 | 29.8 |
| Transmembrane | 522 | 30.8 |
| Repeat | 155 | 9.1 |
| Zinc finger | 22 | 1.3 |
| Coiled coil | 99 | 5.8 |
| Motif | 301 | 17.8 |
| Compositional bias | 512 | 30.2 |
| Active site | 357 | 21.1 |
| Binding site | 520 | 30.7 |
| Non-standard residue | 0 | 0 |
| Modified residue | 878 | 51.8 |
| Lipidation | 115 | 6.8 |
| Glycosylation | 936 | 55.3 |
| Disulfide bond | 935 | 55.2 |
| Cross link | 117 | 6.9 |
| Helix | 1,166 | 68.8 |
| Turn | 954 | 56.3 |
| Beta strand | 1,127 | 66.5 |
| Protein mass (SD), Da | 56,736.5 | 60,773.0 |
| Protein length (SD), residues | 512.5 | 563.3 |
| Number of isoforms (SD) | 2.3 | 1.9 |

### eTable 2: Top ten most annotated terms under each Gene Ontology (GO) category for 1,694 overlapping proteins

| **GO term** | **Frequency** | | **Percentage** |
| --- | --- | --- | --- |
| **Biological process** |  |  | |
| signal_transduction_go_0007165 | 214 | 12.6 | |
| cell_adhesion_go_0007155 | 165 | 9.7 | |
| positive_regulation_of_cell_population_proliferation_go_0008284 | 148 | 8.7 | |
| proteolysis_go_0006508 | 138 | 8.1 | |
| inflammatory_response_go_0006954 | 135 | 8.0 | |
| immune_response_go_0006955 | 126 | 7.4 | |
| positive_regulation_of_transcription_by_rna_polymerase_ii_go_0045944 | 106 | 6.3 | |
| negative_regulation_of_apoptotic_process_go_0043066 | 103 | 6.1 | |
| positive_regulation_of_gene_expression_go_0010628 | 101 | 6.0 | |
| innate_immune_response_go_0045087 | 98 | 5.8 | |
| **Cellular component** |  |  | |
| plasma_membrane_go_0005886 | 692 | 40.9 | |
| extracellular_region_go_0005576 | 635 | 37.5 | |
| extracellular_space_go_0005615 | 581 | 34.3 | |
| cytosol_go_0005829 | 511 | 30.2 | |
| extracellular_exosome_go_0070062 | 469 | 27.7 | |
| cytoplasm_go_0005737 | 461 | 27.2 | |
| membrane_go_0016020 | 328 | 19.4 | |
| nucleus_go_0005634 | 298 | 17.6 | |
| nucleoplasm_go_0005654 | 228 | 13.5 | |
| cell_surface_go_0009986 | 222 | 13.1 | |
| **Molecular function** |  |  | |
| identical_protein_binding_go_0042802 | 266 | 15.7 | |
| calcium_ion_binding_go_0005509 | 169 | 10.0 | |
| metal_ion_binding_go_0046872 | 140 | 8.3 | |
| signaling_receptor_binding_go_0005102 | 136 | 8.0 | |
| protein_homodimerization_activity_go_0042803 | 126 | 7.4 | |
| atp_binding_go_0005524 | 101 | 6.0 | |
| growth_factor_activity_go_0008083 | 95 | 5.6 | |
| cytokine_activity_go_0005125 | 87 | 5.1 | |
| integrin_binding_go_0005178 | 84 | 5.0 | |
| rna_binding_go_0003723 | 82 | 4.8 | |

### eTable 3: Number of proteins by their number of matched SOMAmers and colocalisation results (ANML)

| **No. matched SOMAmers** | **No. proteins** | **No. proteins with colocalising *cis*-pQTLs between OLINK and SomaScan** | | | | | |
| --- | --- | --- | --- | --- | --- | --- | --- |
|  |  | **Total** | **Coloc with 1 SOMAmer** | **Coloc with 2 SOMAmers** | **Coloc with 3 SOMAmers** | **Coloc with 4 SOMAmers** | **Coloc with 6 SOMAmers** |
| **2** | 399 | 152 | 68 | 84 | - | - | - |
| **3** | 56 | 18 | 4 | 9 | 5 | - | - |
| **4** | 11 | 9 | 2 | 0 | 1 | 6 | - |
| **5** | 1 | 0 | 0 | 0 | 0 | 0 | - |
| **6** | 2 | 1 | 1 | 0 | 0 | 0 | 0 |
| **8** | 1 | 1 | 0 | 0 | 0 | 0 | 1 |
| **9** | 2 | 0 | 0 | 0 | 0 | 0 | 0 |

### eTable 4: Number of proteins by their number of matched SOMAmers and colocalisation results (non-ANML)

| **No. matched SOMAmers** | **No. proteins** | **No. proteins with colocalising *cis*-pQTLs between OLINK and SomaScan** | | | | | |
| --- | --- | --- | --- | --- | --- | --- | --- |
|  |  | **Total** | **Coloc with 1 SOMAmer** | **Coloc with 2 SOMAmers** | **Coloc with 3 SOMAmers** | **Coloc with 4 SOMAmers** | **Coloc with 6 SOMAmers** |
| **2** | 399 | 146 | 73 | 73 | - | - | - |
| **3** | 56 | 18 | 5 | 8 | 5 | - | - |
| **4** | 11 | 9 | 2 | 1 | 0 | 6 | - |
| **5** | 1 | 0 | 0 | 0 | 0 | 0 | - |
| **6** | 2 | 0 | 0 | 0 | 0 | 0 | 0 |
| **8** | 0 | 0 | 0 | 0 | 0 | 0 | 0 |
| **9** | 2 | 0 | 0 | 0 | 0 | 0 | 0 |
